# Towards adaptive deep brain stimulation: clinical and technical notes on a novel commercial device for chronic brain sensing

**DOI:** 10.1101/2021.03.10.21251638

**Authors:** Yohann Thenaisie, Chiara Palmisano, Andrea Canessa, Bart J. Keulen, Philipp Capetian, Mayte Castro Jiménez, Julien F. Bally, Elena Manferlotti, Laura Beccaria, Rodi Zutt, Grégoire Courtine, Jocelyne Bloch, Niels A. van der Gaag, Carel F. Hoffmann, Eduardo Martin Moraud, Ioannis U. Isaias, M. Fiorella Contarinoe

## Abstract

**Background:** Technical advances in deep brain stimulation (DBS) are crucial to improve therapeutic efficacy and battery life. A prerogative of new devices is the recording and processing of a given input signal to instruct the delivery of stimulation.

**Objective:** We studied the advances and pitfalls of one of the first commercially available devices capable of recording brain local field potentials (LFP) from the implanted DBS leads, chronically and during stimulation.

**Methods:** We collected clinical and neurophysiological data of the first 20 patients (14 with Parkinson’s disease [PD], five with various types of dystonia, one with chronic pain) that received the Percept™ PC in our centers. We also performed tests in a saline bath to validate the recordings quality.

**Results:** The Percept PC reliably recorded the LFP of the implanted site, wirelessly and in real time. We recorded the most promising clinically useful biomarkers for PD and dystonia (beta and theta oscillations) with and without stimulation. Critical aspects of the system are presently related to contact selection, artefact detection, data loss, and synchronization with other devices. Furthermore, we provide an open-source code to facilitate export and analysis of data.

**Conclusion:** New technologies will soon allow closed-loop neuromodulation therapies, capable of adapting the stimulation based on real-time symptom-specific and task-dependent input signals. However, technical aspects need to be considered to ensure clean synchronized recordings. The critical use by a growing number of DBS experts will alert new users about the currently observed shortcomings and inform on how to overcome them.

## INTRODUCTION

Deep brain stimulation (DBS) is a common practice for the symptomatic treatment of many neurological conditions (e.g., Parkinson’s disease (PD), dystonia, essential tremor).^1,2^ Despite impressive technological and surgical advances over the past 30 years, stimulation therapies are still restricted to continuous stimulation protocols that are tuned manually during in-clinic visits. This lack of adaptability fails to address essential disease-, medication-, and activity-related fluctuations of the clinical condition. To address these limitations, next-generation neurotechnologies are being developed to operate in a closed-loop^3-5^. These devices offer the possibility of automatically adapting stimulation parameters (amplitude, frequency) in response to feedback signals, chronically and in real time. Emerging evidence suggests that adaptive stimulation approaches may exhibit greater efficacy with fewer adverse effects^6-8^. However, translation of such strategies into everyday life is yet to be achieved^9^, and strongly relies on the user-friendliness, quality, and robustness of the sensing capabilities endowed in chronic devices.

Recently, new implantable devices capable of chronically recording local field potentials (LFP) during stimulation became available.^10,11^ Their sensing capabilities should enable better understanding of disease-related brain activity patterns, their evolution over time, and their modulation in response to therapies, bringing the implementation of adaptive stimulation therapies closer to clinical practice.

Here, we report the potential applications and pitfalls that emerged when using Percept™ PC (Medtronic PLC, USA) in the first 19 patients implanted at our centers. We provide relevant surgical, technical, and operational aspects to be considered to maximize its performance and signal quality. We also describe tests performed in a saline bath to validate recording quality.

## METHODS

### Patients

We collected clinical and neurophysiological data for the first 19 patients (14 PD implanted in the STN, five dystonia implanted in the GPi; 11 males) that received the Percept PC at our three centers: Lausanne University Hospital (CHUV), University Hospital of Würzburg (UKW), and Haga Teaching Hospital/Leiden University Medical Center (Haga/LUMC). Patients were not selected based on specific characteristics and the implant was performed in the context of clinical practice.

The severity of PD and dystonia symptoms was assessed using clinical rating scales by an experienced movement disorders clinician, as part of the clinical routine (Table 1).

**Table 1:** Demographic information and Percept™ PC recording modes available for analysis

| Subject<sup>a</sup> | Diagnosis | Age at surgery (years range<sup>b</sup>) | Disease duration at recording (years) | Clinical score (OFF/ON)<sup>c</sup> | Preoperative medication<sup>d</sup> | Time of recordings since lead implantation | Survey |  | Setup | Timeline | Events | Streaming |  |
| --- | --- | --- | --- | --- | --- | --- | --- | --- | --- | --- | --- | --- | --- |
|  |  |  |  |  |  |  | Survey | Indefinite streaming |  |  |  | Stimulation Off | Stimulation On |
| **CH1** | PD | 65-70 | 13 | 37/18 | 1150 | 6d, 3m |  | OFF, ON |  |  |  |  |  |
| **CH2** | PD | 70-75 | 8 | 31/9 | 1670 | 5-6d |  | ON |  |  |  |  |  |
| **CH3** | PD | 45-50 | 4 | 16/7* | 1175 | 2d |  | ON |  |  |  |  |  |
| **CH4** | PD | 40-45 | 5 | 63/12* | 500 | 6d |  | ON |  |  |  |  |  |
| **CH5** | PD | 60-65 | 2 | 45/33 | 1350 | 2d-3m |  |  | OFF |  |  |  | OFF<sup>e,f</sup> |
| **CH6** | PD | 70-75 | 13 | 30/9 | 1600 | 3-5d |  | ON | ON | ON | ON |  | ON<sup>e</sup> |
| **CH7** | PD | 55-60 | 7 | 50/30 | 475 | 3-5d |  | ON | ON |  |  |  | ON<sup>e</sup> |
| **CH8** | PD | 50-55 | 4 | 63/43 | 1617 | 3d |  | ON | ON |  |  |  | ON<sup>e</sup> |
| **CH9** | PD | 65-70 | 14 | 26/8 | 1325 | 5d |  | OFF | OFF |  |  |  | OFF<sup>e</sup> |
| **NL1** | PD | 60-65 | 6 | 38/15 | 1000 | 2d-8m | OFF, ON |  | OFF, ON | ON |  | OFF | OFF, ON<sup>e</sup> |
| **NL2** | PD | 75-80 | 4 | 34/19 | 600 | 10d-7m | OFF |  | OFF, ON | ON |  | OFF | OFF, ON<sup>e</sup> |
| **PW1** | PD | 65-70 | 24 | 38/8 | 1100 | 8y, 6m<sup>g</sup> | OFF |  | OFF |  |  | OFF, ON | OFF, ON |
| **PW2** | PD | 70-75 | 16 | 38/13 | 610 | 5y, 4m<sup>g</sup> |  |  |  |  |  | OFF |  |
| **PW3** | PD | 50-55 | 17 | 55/4 | 1300 | 5y, 7m<sup>g</sup> |  |  |  |  |  | OFF |  |
| **PW4** | CD | 35-40 | 35 | 16-4 and 16-0-2.5 | - | 8y, 11m<sup>g</sup> |  |  |  |  |  | X |  |
| **PW5** | MD | 60-65 | Child onset | 5-6 and 17-9-7.5 | - | 3m, 11d | X | X | X |  |  | X |  |
| **PW6** | MD | 30-35 | Child onset | 50.5-6 and 20-8-13 | - | 1m, 18d | X | X | X |  |  | X |  |
| **PW1A** | CD | 45-50 | 12 | 8-3 and 18-13-8,75 | - | 4d | X | X |  |  |  |  |  |
| **PW8** | CD | 60-65 | 15 | 5-2 and 20-04-9.5 | Trihexyphenidyl 1 mg TID | 5d | X | X | X |  |  | X |  |

<sup>a</sup> Subjects indicated with code CH were studied at the Lausanne University Hospital (CHUV, Switzerland); subjects with code NL at the Haga Teaching Hospital/Leiden University Medical Center (Haga/LUMC, Netherlands); subjects with code PW at the University Hospital of Würzburg (UKW, Germany).
<sup>b</sup> Year range is provided instead of precise age to reduce the chance of identification
<sup>c</sup> Here and in the rest of the table "OFF" and "ON" refer to the medication status, unless otherwise indicated. Medication status is labeled ON if the patient received any anti-Parkinsonian/anti-dystonic drugs within 12h before the recording, otherwise labeled as OFF. Clinical scores for Parkinson's disease patients are MDS-UPDRS or UPDRS (labeled \*), measured pre-operatively OFF medication and 1h after medication intake. Clinical scores for dystonia measured preoperatively are Burke-Fahn-Marsden Dystonia Rating Scale (severity - disability) and Toronto Western Spasmodic Torticollis Rating Scale (TWSTRS) (severity - disability - pain).
<sup>d</sup> Anti-Parkinson medication is expressed in LEDD mg equivalent (according to Tomlinson CL, Stowe R, Patel S, Rick C, Gray R, Clarke CE. Systematic review of levodopa dose equivalency reporting in Parkinson's disease. Mov Disord. 2010 Nov 15;25(15):2649-53. doi: 10.1002/mds.23429. PMID: 21069833).
<sup>e</sup> The recordings were performed "on" stimulation, but the stimulation amplitude was at 0 mA except during synchronization bursts.
<sup>f</sup> Streaming performed during contact review and stimulation titration.
<sup>g</sup> These patients received the Percept PC as a replacement for the Activa PC due to battery consumption.
Abbreviations: CD, Cervical Dystonia; CP, Chronic Pain; d, days; F, female; M, male; m, months; MD, Myoclonus dystonia; MDS-UPDRS, Movement Disorders Society-Unified Parkinson's Disease Rating Scale; n.a., not available; PD; Parkinson's disease; y, years.

### Surgical procedure

Four patients received Percept PC during replacement of their implantable pulse generator (IPG). All others received it simultaneously to lead implantation or five days afterwards (two patients). In one patient (PW4), the IPG was implanted in the right abdominal region; in all others it was implanted in the chest (left: NL1, NL2, PW2-3; right: CH1-9, PW1, 5-8, PW1A).

All patients received bilateral leads (3389, Medtronic, PLC, USA) with four cylindrical contacts – for clarity, subsequently named 0-3L and 0-3R.

The surgical procedure of each center has been previously reported^12-15^. No specific procedures were followed for Percept PC implantation. (Supplementary file 1)

### Recordings

The Percept PC can continuously record LFP in real time also during active stimulation, and to transmit them wirelessly. The device uses a sampling frequency of 250Hz, and contains two low-pass filters at 100Hz and two high-pass filters at 1Hz, and 1Hz or 10Hz (user defined)^16^. Bipolar recordings can be performed in several modes (BrainSense™, Table 2). Data can be visualized online (Fig.1), saved, and exported in a JavaScript Object Notation (JSON)-format file. (Fig.2) The software to analyze the JSON file is not provided and must be built in-house.

**Table 2.** Main functions and features of the available recording modes with the Percept™ PC device.

|  |  |  |
| --- | --- | --- |
| BrainSense™<br>*Survey* | *Survey*<br>(In-clinic use) | – Recordings from all possible contact pairs (Fig.1A).<br>– Impedance check and artefact screening performed.<br>– Measurement duration 90 s.<br>– The user interface (tablet) displays local field potential (LFP) magnitude [micro volts peak, $\mu\text{vp}$] vs. frequency [Hz] (about 21s of data for each pair).<br>– Time domain data available in the .json file.<br>– Stimulation is "off" during the measurement. |
|  | *Survey Indefinite Streaming*<br>(In-clinic use) | – Available on the same screen of the *Survey*.<br>– Stream as long as desired from all stimulation-compatible contact pairs (e.g., 0-3, 1-3, 0-2)<br>– No user interface output on the recorded data.<br>– Time domain data available in the .json file.<br>– Stimulation is "off" during the measurement. |
| BrainSense™<br>*Streaming* | *Setup*<br>(In-clinic use) | – Impedance check and artefact screening are performed.<br>– The artefact screening can be turned off.<br>– Measurement duration 90 s.<br>– Data in the frequency domain (LFP magnitude [$\mu\text{Vp}$] vs. frequency [Hz]) are visible online.<br>– The tablet indicates the largest peak in the beta or gamma frequency range per contact pair (only for stimulation compatible pairs and only if $>1.1\mu\text{Vp}$).<br>– Time domain data available in the .json file.<br>– The user can manually select a contact pair and a frequency band for subsequent recordings.<br>– Stimulation is "off" during the measurement. |
|  | *Streaming*<br>(In-clinic use) | – Must be preceded by LFP sensing setup (BrainSense™ Setup).<br>– Online recordings from a single pre-defined contact pair for each hemisphere (if both have a complete Setup)<br>• The tablet displays the selected power and current in real time and over the entire recording (Fig.1B).<br>– Time domain data available in the JSON file.<br>– Stimulation can be "off" or "on". |
|  | *Timeline*<br>(at-home use) | – Enabled by selecting "Passive" in *BrainSense Streaming*(Figure 1B)<br>– LFP data are chronically collected (up to 60 days)<br>– Measures 250Hz time domain data converted to the frequency domain<br>– Recording of the power of the pre-selected frequency band (selected frequency $\pm 2,5\text{Hz}$) in the preselected contact pairs (depending on active stimulation contact)<br>– Computes 10min-averaged power |
|  | *Events* | – Patients can manually mark events (e.g., fall, took |
|  | (at-home use) | medication, on time, dyskinesia) during Timeline recordings using the patient programmer.<br>• LFP are recorded for 30s directly after trigger.<br>– The average power spectrum density (PSD) estimate of each event is stored and can be retrieved in clinic. |

**Figure 1.**
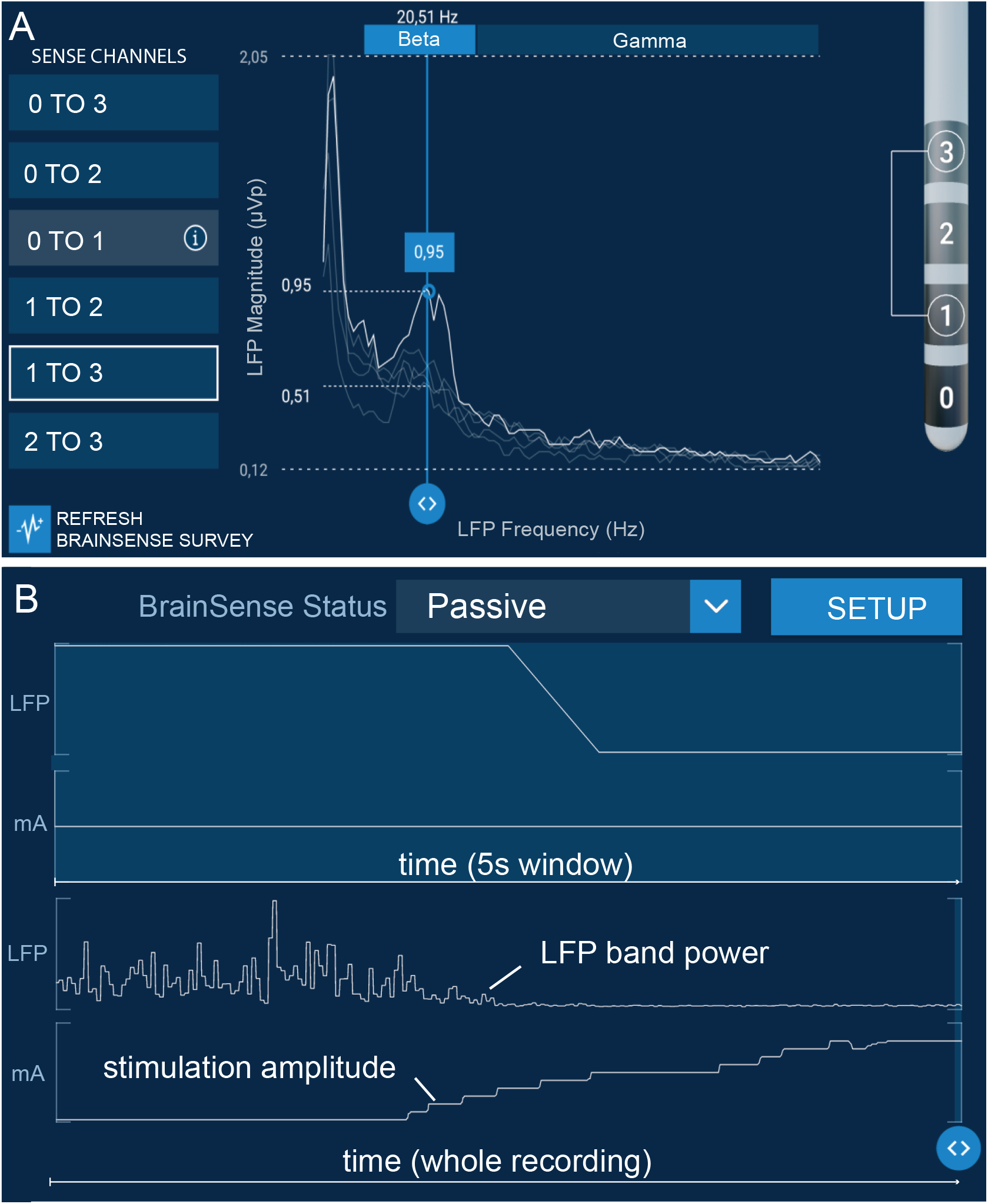
Online evaluations using BrainSense^™^. **(A)** Screenshot of the Percept^**™**^ PC User Interface (tablet) displaying the power spectrum density (PSD) generated in the BrainSense *Survey* modality (patient NL1, left lead, meds-off condition). The highest beta power could be identified between contacts 1 and 3. Monopolar stimulation from contact point 2 had the best effect but induced dyskinesia, thus contact 1 was chosen for chronic stimulation. **(B)** Online visualization of LFP power sensing for a pre-selected frequency band in the BrainSense *Streaming* mode (one new datapoint visualized every 500ms), while manually increasing stimulation amplitude (patient CH5, right subthalamic nucleus). Increase in stimulation amplitude induced a decrease in beta-band power after reaching 2mA. Concurrent clinical motor evaluations confirmed improvements in rigidity associated with LFP beta power modulations.

**Figure 2.**
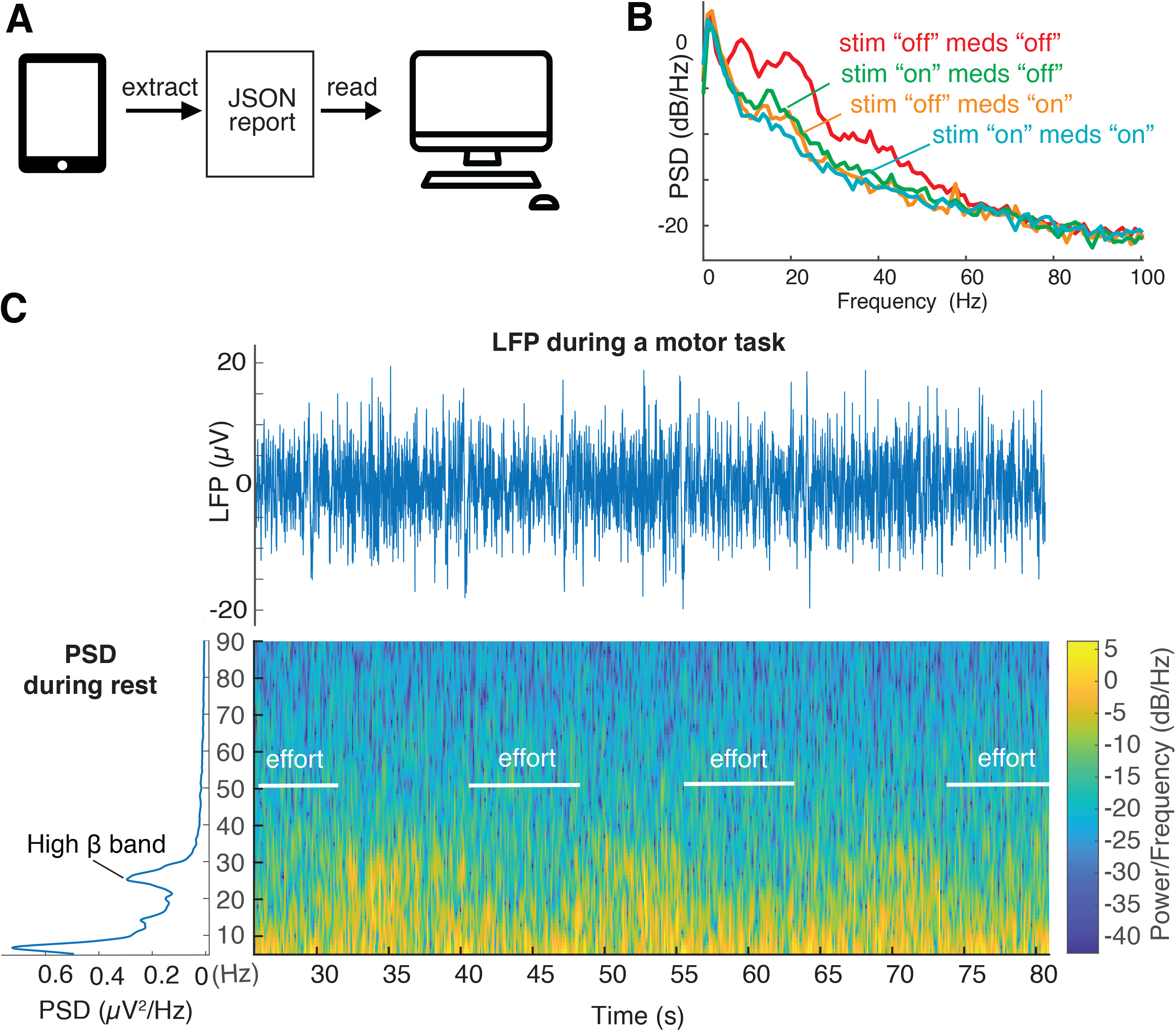
Offline evaluations of high-resolution local field potential (LFP) recordings modulated by medication, stimulation, and movement. **(A)** Raw high-resolution (250Hz) LFP signals can be extracted and processed offline for further analysis, complementary to the online visualization. **(B)** Power spectrum density (PSD) estimates of resting state recordings (patient PW3, sitting with eyes open) in four different conditions: meds “off” (overnight suspension of all dopaminergic drugs.), meds “on” only (1h after intake of a standard levodopa dose), stim “on” only (stimulation with optimal amplitude, 180Hz), and combined stim “on” and meds “on”. Suppression in beta power is captured both in the stimulation-only and medication-only conditions, and with the cumulative effect of both medication and stimulation. **(C)** Raw LFP signal and Welch’s PSD estimates of BrainSense *Survey Indefinite Streaming* recordings at rest and spectral changes over time during iterative movements (knee extension movements while sitting, patient CH6, right STN, contacts 0-2, meds “off” and stim “off”). Unprocessed LFP and spectrogram accurately captured expected movement-related changes in beta power.

### Patients recordings

Patient recording details are reported in Table 1 and Supplementary file 1. Recordings were performed *in* the eyes-open resting state and, in some patients, during a motor task (isometric knee extension or walking) or unperturbed walking^14,17,18^. In three patients, *Timeline* recordings were obtained.

### *In vitro* recordings

A DBS lead (3389) was inserted in a saline bath and simultaneously connected to the Percept PC and to a high-resolution external amplifier able to record at 24414.06Hz (RZ5D, Tucker Davis Technologies, TDT, USA). Signals from both systems were synchronized by sending an external 10Hz sinusoidal signal generated with Agilent 33210A LXI at 100mA for a few seconds. Tests aimed to: (i) validate the nominal sampling frequency of the Percept PC; (ii) evaluate stimulation artefacts; (iii) validate synchronization methods.

### Data analysis

All LFP recordings were exported from the Percept PC tablet as JSON files, imported into MATLAB (Mathworks, Natick, MA) with a custom-built toolbox available at https://github.com/YohannThenaisie/PerceptToolbox.git, and analyzed using custom-built code.

For the removal of the cardiac artefact, we computed the singular value decomposition of LFP data epoched around the QRS peaks. The component resembling the QRS complex was visually identified (namely the ones explaining more than 97.5% of the variance) and subtracted from the raw data^18^.

### Analysis of the recordings in PD patients

Beta-band analysis was performed on *Survey Indefinite Streaming* or *Survey* data (Supplementary Table 1, Supplementary file 1, Fig.3). For each *Survey Indefinite Streaming*, we reconstructed bipolar LFP signals from adjacent contacts by subtracting channels similarly to standard EEG montages. Power spectrum density (PSD) estimates were computed via Welch’s method (*pwelch* function). We defined the frequency of the beta peak f_Peak_ as the frequency with the local maxima power in the 13-35Hz range. We visually verified each PSD for the presence of beta (13-35Hz) and gamma (60-90Hz) bands. For each subthalamic nucleus (STN), we reported the contact pair with the highest f_Peak_ power in the beta band. Short-time Fourier transform was applied on raw LFP from all *Streaming* recordings (Table 1).

**Figure 3.**
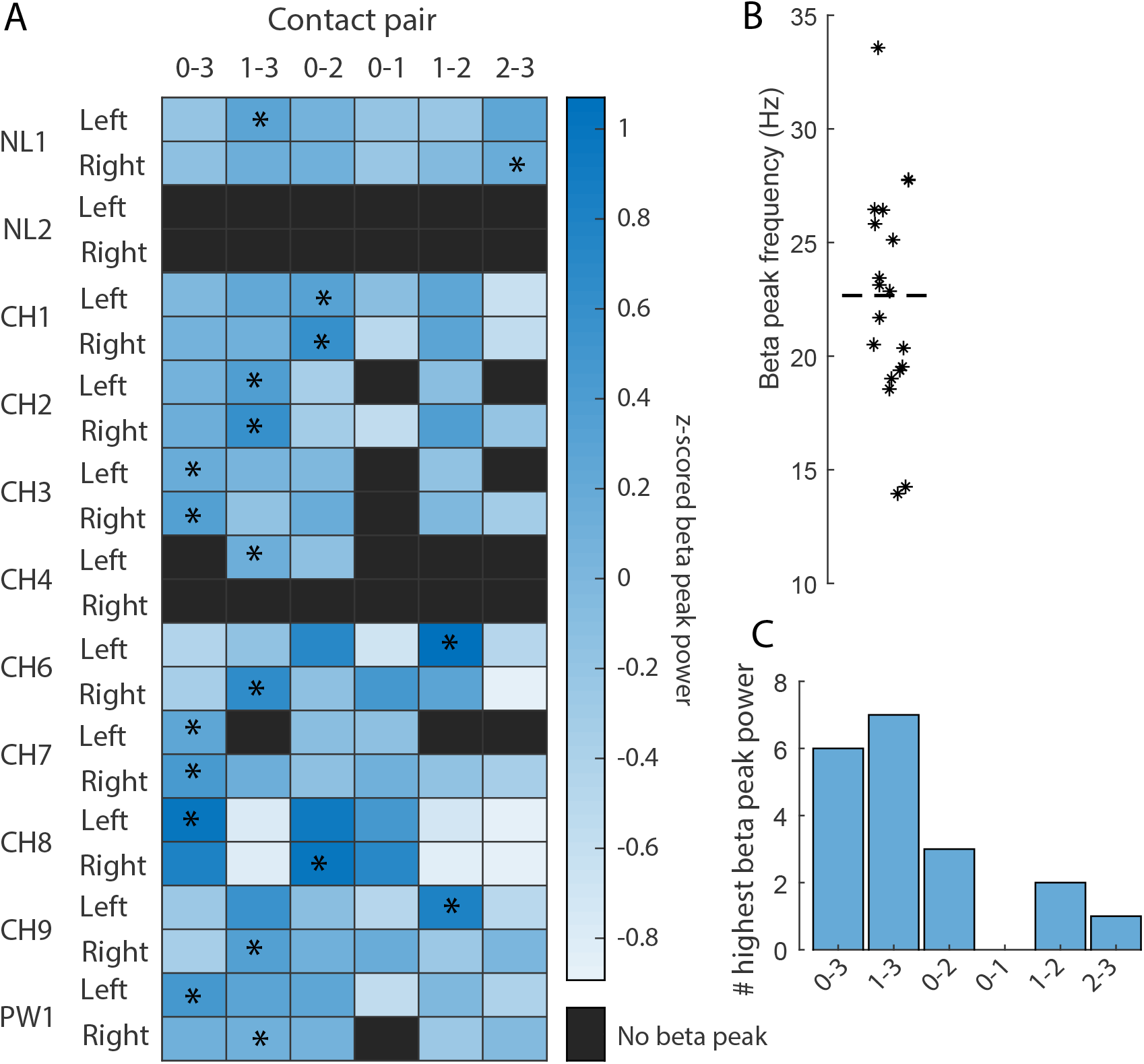
Beta peak frequency and power in all contact pairs of 22 subthalamic nuclei (STN) recorded with *Survey* and *Survey Indefinite Streaming* modalities during resting. **(A)** Power of the beta peak in all contact pairs of 22 STN, normalized by STN. A contact pair is displayed as black when no peak could be identified in the beta range (13-35Hz). For each STN, the contact pair with the maximum beta peak power is labeled with a star. **(B)** Peak frequency of the contact pair with the highest beta power for the 19 STN in which the beta peak was identified in at least one contact pair. The horizontal dashed line indicates the average beta frequency of all STN. **(C)** Number of times each of the contact pairs was identified as the one with maximum beta power across the 19 STN in which a beta peak was present.

### Analysis of the recordings in dystonic patients

Theta-band analysis was performed on *Survey* data. We reconstructed bipolar LFP signals from adjacent contacts as for PD patients. PSD was computed using Welch’s method and 1/f component removal^19^.

For each contact pair, PSD was normalized for the standard deviation computed between 6-96Hz for comparison across patients,^20^ visually inspected for peaks in the theta band (4-12Hz) (Fig.4), and checked for movement artefacts (Supplementary Fig.1). LFP recordings during gait (PW4) were synchronized with the kinematic data (Fig.5). Body kinematics were measured with a full-body marker set and a motion capture system (SMART-DX, BTS, Italy)^21,22^.

**Figure 4.**
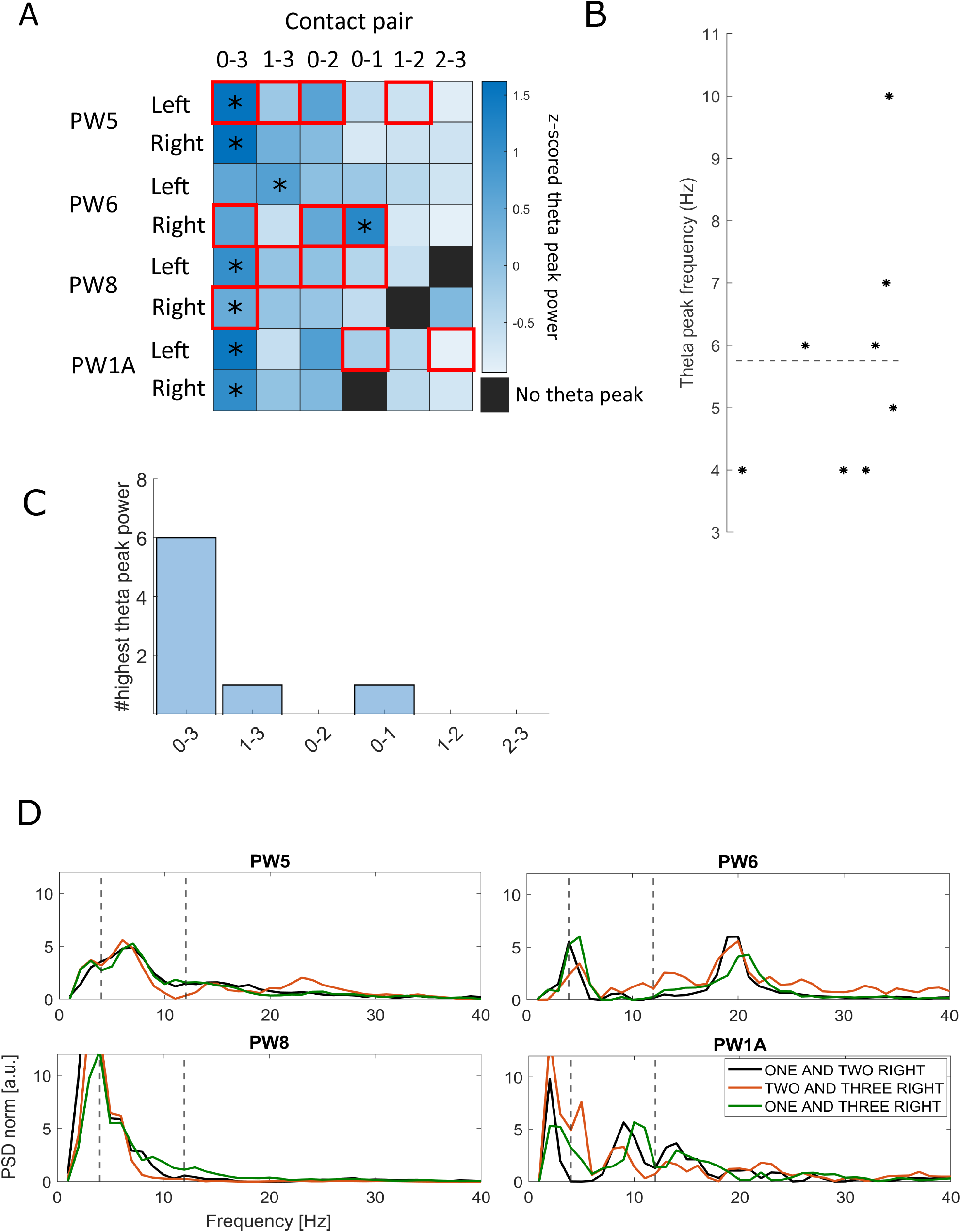
Theta peak frequency and power in all contact pairs of eight globus pallidus internus (GPi) recorded using the *Survey* mode during resting. **(A)** Power of the theta peak in all contact pairs of eight GPi nuclei, normalized by GPi. A contact pair is displayed as black when no peak could be identified in the theta range (4-12Hz). For each GPi, the contact pair with the maximum theta peak power is labeled with a star. Red boxes indicate contact pairs labelled as artefactual by the device. **(B)** Peak frequency of the contact pair with the highest theta power for all GPi. The horizontal dashed line indicates the average theta peak frequency of all GPi. **(C)** Number of times each of the contact pairs was identified as the one with maximum theta power across the eight GPi. **(D)** Power spectrum density (PSD) of local field recordings (LFP) recorded by contact pairs labeled as non-artefactual in all patients. Vertical dashed lines indicate the theta band. Of note, a peak in the theta band was evident in patients PW5, PW6, and PW1A from all recording pairs. Patient PW6 also showed a prominent peak in the beta band. Patient PW8 did not show a clear modulation in either the beta or theta bands, apart from a peak at 4Hz recorded from the electrodes pair 1-3R.

**Figure 5.**
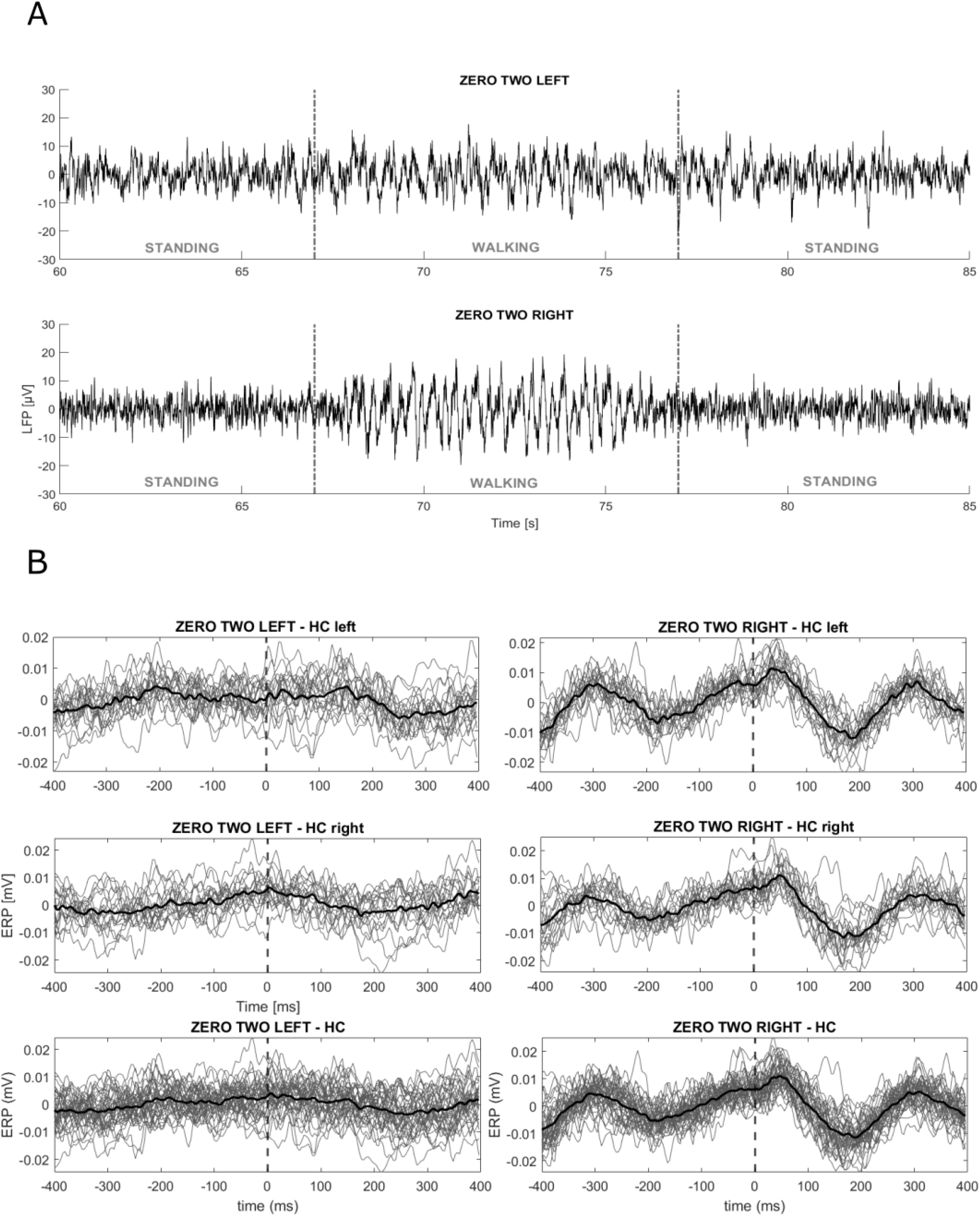
Movement-related artefacts during walking. One dystonic patient (PW4) was asked to stand quietly and to start walking over an 8 m long walkway after a verbal cue. The task was repeated four times. Body kinematics were measured with a full-body marker set and a motion capture system (SMART-DX, BTS, Italy). **(A)** Raw data of local field potential (LFP) recordings in *Streaming* mode in stimulation “off” from contact pairs 0-2L and 0-2R. The vertical grey dashed lines identify the walking window (from about 67 s to 77 s), preceded and followed by quiet standing. Movement-related artefacts are visible in the raw signal. **(B)** LFP of all walking trials epoched in 800-ms windows centered at the heel contacts (vertical dashed lines), as detected by the kinematic assessment. In total, 37 epochs of gait were analyzed. The left and right columns show the data of the left and right hemispheres epoched with respect to the left (first line) and right (second line) heel contacts (HC), and to all the heel contacts (third line). In each subpanel, the grey lines represent the LFP recorded in each epoch and the thick black line represents the average across all epochs. We found a modulation on the right hemisphere only, not related to the side of the ongoing stepping.

### Ethics

The Medical ethical committee Leiden Den Haag Delft, the Ethik-Kommission of the University Hospital Würzburg and the Ethical Committee of the Canton of Vaud (CER-VD) approved the respective studies and/or waived review for the data collection of the respective center.

All patients gave written informed consent according to the Declaration of Helsinki.

## RESULTS

### 1. Outcomes of clinical relevance

#### Recordings of STN beta band (13-35Hz) in PD

A beta peak was identified in 19 out of 22 STN for whom the *Survey or Survey Indefinite Streaming* mode was available, at an average frequency of 22.6Hz (SD ±4.9Hz) (Fig.3A-B). In three STN, no contact pair displayed a beta band. In 13 out of 19 STN, the maximum beta peak was found in contact pair 1-3 or 0-3 (Fig.3). In all but three STN (of three different patients), the clinically chosen contact for chronic stimulation was either in between or one of the contact pairs displaying the maximum beta peak (Supplementary Table 1). In some patients, it was also possible to identify a stimulation amplitude-dependent modulation of the beta power (Fig.1B, Supplementary Fig.2). Finally, we were able to confirm that the Percept PC captures modulations in beta power induced by STN-DBS and levodopa at rest (Fig.2B) and arising with movements during a motor task (Fig.2C).

#### Recordings in dystonic patients

A theta peak was identified in all eight globus pallidus internus (GPi) nuclei, at an average frequency of 5.7Hz (SD ±2.1Hz) (Fig.4). In six out of eight GPi, the maximum theta peak was found on contact pair 0-In these recordings, about 27% of the contact pairs were labeled as artefactual by the Percept PC (see below). Consecutive multiple *Survey* recordings showed high variability of LFP measurements. The signal recorded in the same patient (PW05) by the same contacts during consecutive sessions was differently identified as artefactual or non-artefactual (Supplementary Fig.1), possibly because of the influence of the episodic movement artefacts.

#### At-home recordings

In the three patients (NL1, NL2, and CH6) recorded with *Timeline*, we observed daily and circadian beta power fluctuations. One patient (CH6) was asked to mark events of freezing of gait (Fig.6).

**Figure 6.**
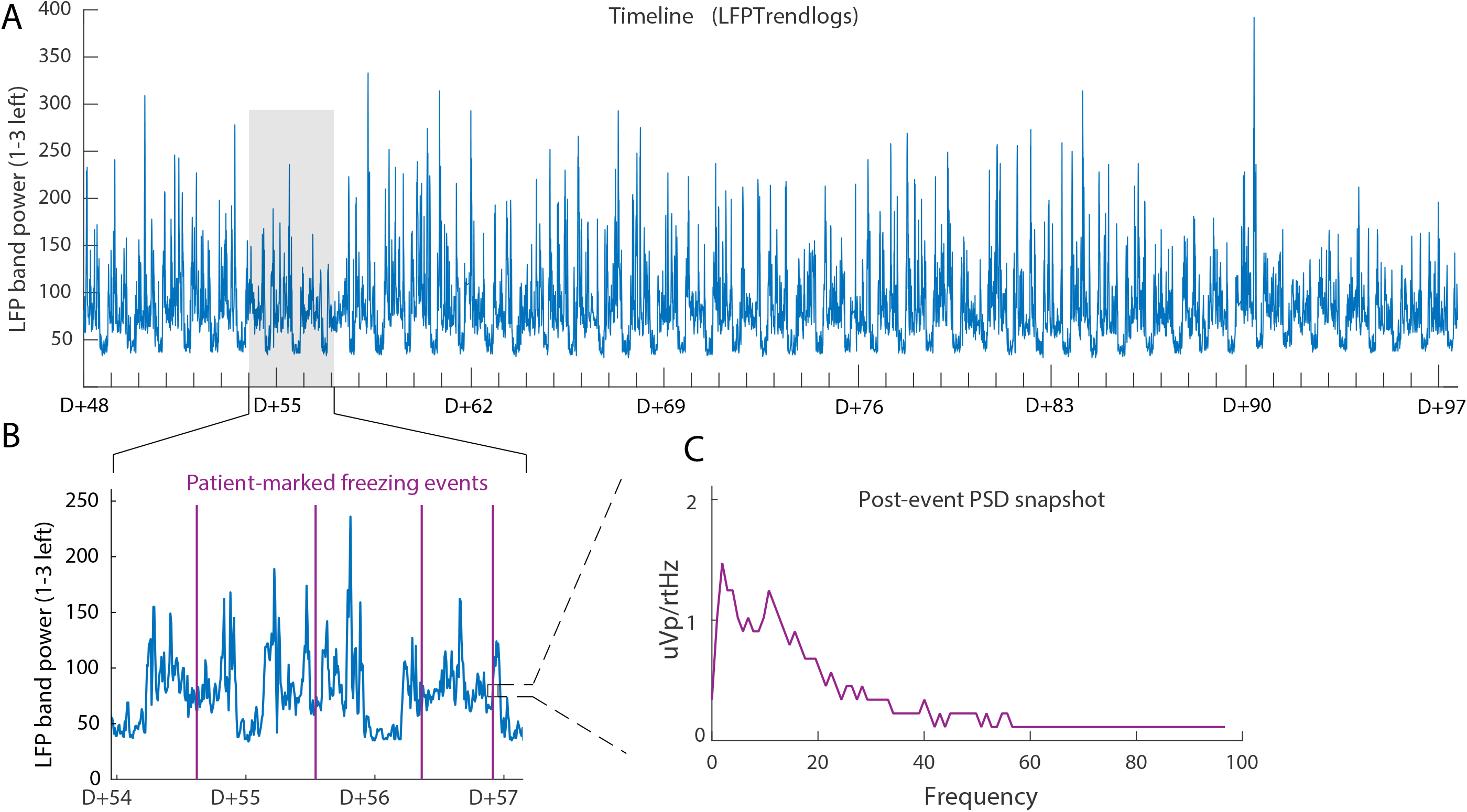
At-home recordings and marked events. In patient CH6, at-home recordings were performed with the *Timeline* modality. A frequency band in the beta range (23.39Hz) was selected by the clinician to be chronically monitored in “passive mode” by contact pair 1-3L. Stimulation was “on” on contact 2L at the optimal therapeutic value (2.9-3.2 mA). **(A)** Local field potential (LFP) band power between 48 days and 97 days after surgery. **(B)** 3-day close-up view of the recording in (A). Signal quality was appropriate to capture circadian and daily fluctuations in power that arose over time, including power depressions at night. In addition, the patient manually reported freezing of gait events (purple lines) on their patient controller. **(C)** Example power spectrum density (PSD) of an event reported by the patient (left subthalamic nucleus displayed only). For each marked event, the Percept™ PC computed bilateral PSD estimates over a ∼30 s window following each marker. This data is only proposed as a representation of this function (see text).

### 2. Technical evaluations

#### Sampling frequency

By recording a pre-defined 10.0Hz sinusoid signal of amplitude 500mV generated by a function generator with Percept PC in a saline bath, we estimated the sampling frequency to be 249.7Hz (over 745 oscillations). This confirms the manufacturer’s nominal value (Fs=250Hz).

### 3. Troubleshooting

Advice and recommendations for successfully recording LFP signals with the Percept PC device are summarized in Table 3.

**Table 3.** General considerations to improve local field potential (LFP) recordings with the Percept™ PC device.

| Tips and Tricks |  |
| --- | --- |
| Recording procedure and data export:<br>*How to avoid data loss* | – Keep the components close to each other<br>• Keep the User Interface (tablet) in a 2-meter range from the Communicator.<br>• Keep the Communicator close to the implanted pulse generator (IPG), e.g., by fixing it on the patient.<br>• Avoid any obstruction between the tablet and the Communicator.<br>– Avoid excessive recording file size<br>• Keep the duration of the total recording session preferably below 10 minutes.<br>• When streaming, close and re-open a new session on the tablet at least every 10 minutes.<br>– During *Streaming* recordings, do not change the frequency of stimulation or the programming group (this would cause a drop in the connection).<br>– While recording, monitor the tablet for gaps in recordings.<br>– For convenience, close the session on the tablet and launch a new one after each recording in order to obtain a single JavaScript Object Notation (JSON) file for each recording (all recordings performed in the same session are saved in the same JSON data extraction file).<br>– Make sure the tablet's time is the current time (connect to internet for time update), update the IPG time, and note down the time of start and end of each recording session to be able to match the JSON file with the session after data export (JSON files are named with a timestamp that corresponds to the date and time of the export from the tablet, and not that of the recording). The correct information can be found in the SessionDate in the JSON file. |
| Synchronization with other devices<br>*Possible methods* | – Switch deep brain stimulation (DBS) "on" and "off" to generate an artefact on the LFP recording, which can be simultaneously sensed using EEG or an electromyography probe.<br>• DBS should be switched on and off abruptly: deactivate the "ramp" option and go straight to the intended amplitude (without allowing the automatic gradual amplitude increase)<br>• For "stim off" recordings, an ad-hoc stimulation program could be created with frequency of 80-90Hz, pulse width of 90 μs, and amplitude as high as tolerated by the patient.<br>• For "stim on" recordings, the usual stimulation parameters will be adopted. |
|  | • Consider repeating the “on” and “off” switch at the end of the recordings.<br>– In case time synchronization with external devices is required, it is important to always check that the duration of the LFP data matches the duration of the recording to identify potential data losses. |
| Artefacts management | – Verify the presence of artefacts (i.e., cardiac and motion artefacts) in the LFP signals by performing *Survey* and *Setup* recordings.<br>• Perform multiple recordings to better identify artefactual contacts in case of episodic artefacts (e.g., dystonic and myoclonic jerks).<br>– If possible, avoid (chronic) recordings from contact pairs labeled as artefactual by the *Survey* and *Setup* mode.<br>– Longer recordings (with the *Survey Indefinite Streaming* or *Streaming* modes) might be more robust to episodic artefacts. In patients with dystonia, we recommend recordings of at least 2 minutes.<br>– For recordings during movements, it is advisable to have synchronized kinematic data and epoch the LFP signal to relevant movement events (e.g., heel contacts during gait). |

#### Size and structure of exported files

Each session may be exported as one JSON file for offline analysis. Importantly, multiple consecutive recordings within the same session are appended and saved in a unique JSON file during export, which make it difficult to later identify the single recordings.

We observed that recordings longer than 10 minutes (performed before December 2020) resulted in export failures and data loss, possibly due to excessively large files. Furthermore there was no correspondence between the User Interface (tablet) recording names and times and the names attributed to the JSON files, which were identified by a timestamp with the date and time of the export from the tablet (and not of the recording). The time of start and end of the recording is saved within the JSON file.

#### Data loss during online streaming

We encountered two situations of temporary loss of data streaming:

- The data stream lost a few data packets, but the recording was not interrupted. Such events can be observed as an interruption of the continuous LFP line displayed on the tablet (Fig.7Ai). We experienced data-streaming loss when the Communicator (positioned onto the IPG) and the Tablet were >3-5m apart, or when an obstacle (including the patient’s body) was in-between. Of note, data appeared as a continuous matrix, without any indication of the missing samples in the JSON file. The missing data packets could not be retrieved. However, the JSON file contained the timestamps of the received data packets (TicksInMses data field) and the number of samples of each data packet (GlobalPacketSizes data field). These two metainformation could be used to infer the time of the missing samples and preserve time synchronization with other devices (see Toolbox).
- The data stream was temporarily interrupted (Fig.7Aii). This can happen when the Communicator and Tablet or IPG are far from each other, or when changing the stimulation frequency or programming group with active *Streaming* mode (Fig.7C). In this case, data recorded after the interruption was stored in a new data matrix after the streaming was retrieved. Although lost data could not be recovered, its duration could be inferred from metainformation (TicksInMses) to preserve alignment with other devices. No data packet was lost when changing the stimulation amplitude (mA) with active *Streaming* mode.

In both cases, the duration of the missing data packets could be inferred using metainformation saved in the JSON file, which allowed realigning the data (Fig.7B). We provide a Matlab Toolbox for this purpose (see Methods).

**Figure 7.**
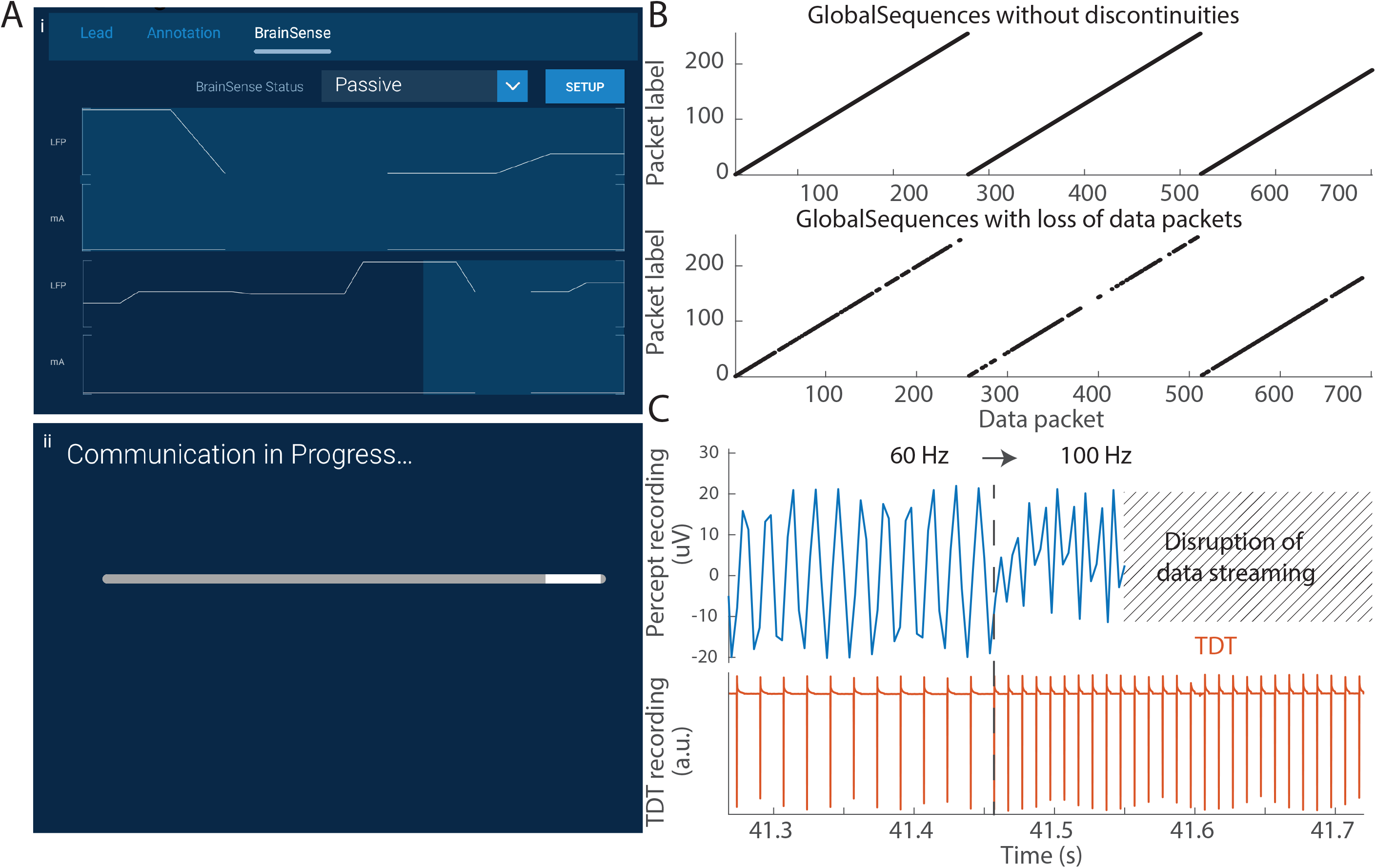
Data-packet loss due to transmission failures, and data re-alignment. **(A)** We encountered two main data transmission problems, which would display either (i) a discontinuity in the data trace display, and resulted in a loss of packets, or (ii) a loading bar indicating communication difficulties, which resulted in the data recording being cut in two matrices. **(B)** The presence and location of missing data packets could be inferred (and corrected) from the exported JavaScript Object Notation (JSON) file. The fields ‘GlobalSequences’, ‘TicksInMses’, and ‘GlobalPacketSizes’ provided the necessary information to ensure a proper alignment in time, even in presence of missing data. **(C)** Changes in the frequency of stimulation temporarily terminated the data transmission, and started a new recording, resulting in data loss. Tests in saline water confirmed that changes from 60Hz to 100Hz during a recording (1.0mA, 60μs) resulted in such disruption in the recording.

#### Artefact detection

##### Stimulation-related artefacts

For stimulation frequencies below the Nyquist frequency (125Hz), a stimulation-related peak artefact corrupted the PSD at the corresponding frequency and its ascending harmonics (Fig.8). For stimulation frequencies above the Nyquist frequency (i.e., 130Hz and above), we also measured stimulation artefacts but at lower frequencies. (Fig.8). This effect is due to aliasing and is explained by: *artefact frequency* = *sampling frequency* – *stimulation frequency*. These in-patient observations were verified in a saline bath setup, by comparing Percept PC signals with recordings performed using a high-resolution amplifier that is not limited to 125Hz Nyquist frequency. (Fig.8C) The amplitude of these artefacts decreased as the stimulation frequency increased from 125Hz upwards. This could be an effect of the 100Hz low-pass filter.

**Figure 8.**
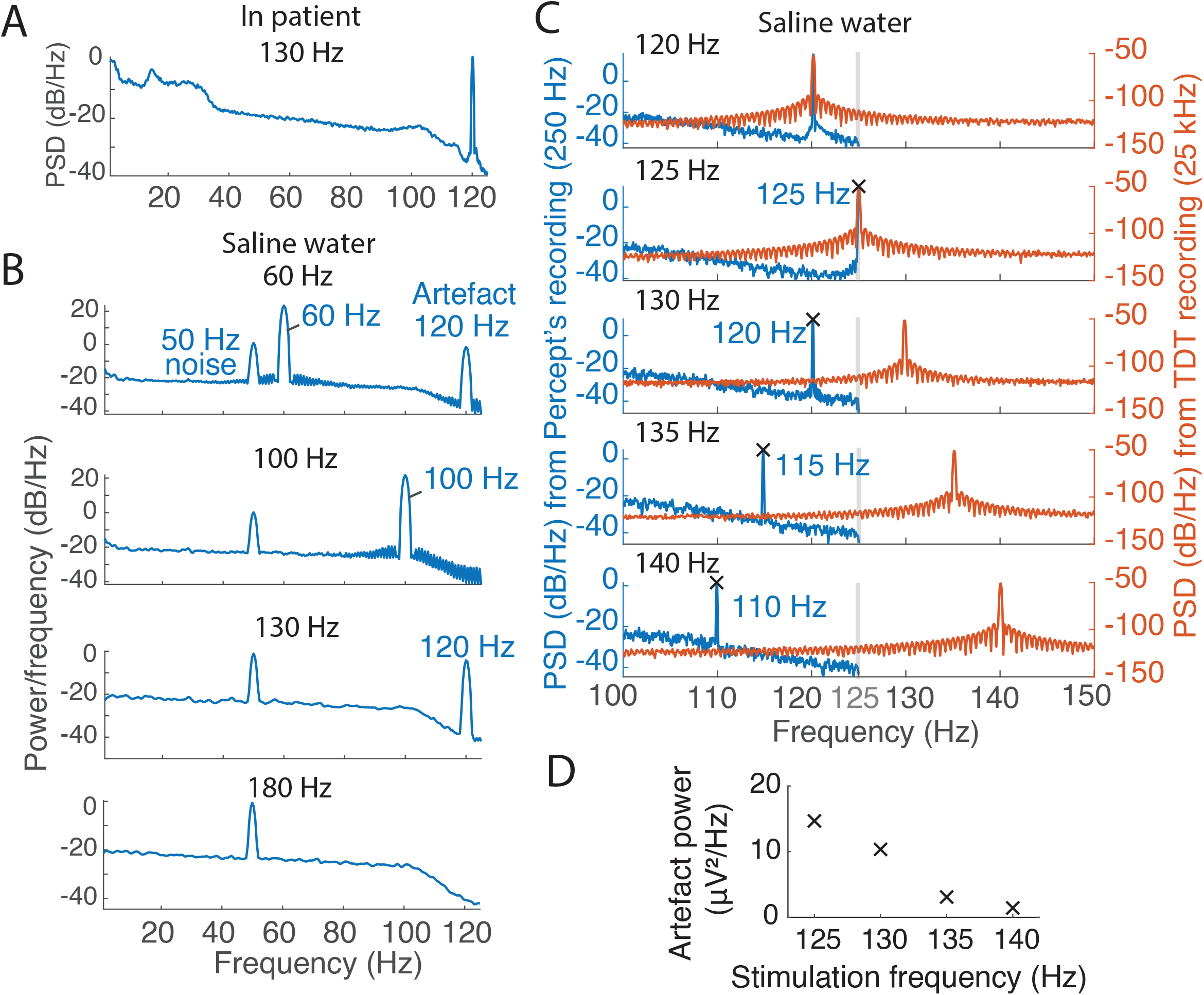
Stimulation aliasing artefacts. **(A)** Power spectrum density (PSD) estimate of local field potential (LFP) signals during a rest recording with contact pair 0-2R with continuous 130Hz deep brain stimulation (1R-C+, 1mA, 60μs) in *Streaming* mode (patient CH6). A power peak corrupted the spectrum at 120 Hz (According to the formula *artefact frequency = sampling frequency (250Hz)– stimulation frequency*). **(B)** PSD estimates in saline water during *Streaming* recordings, for stimulation profiles of four frequencies (60Hz, 100Hz, 130Hz, and 180Hz). Stimulation under the 125Hz-Nyquist frequency (i.e., 60Hz, 100Hz) induced power peaks at the stimulation frequency and potential harmonics. Stimulation at 130Hz induced an artefact at 120Hz. No artefacts were apparent during 180Hz stimulation. **(C)** We stimulated in saline water at various frequencies (1.0mA, 120-140Hz, 60μs) while recording with Percept in *Streaming* mode and with an amplifier of 25kHz sampling frequency. Stimulations above 125Hz induced artefacts at frequencies symmetric to 125Hz in the Streaming recording only. For example, stimulation at 130Hz (5Hz above the Nyquist frequency) induced artefacts at 120Hz (5Hz below). **(D)** In saline water, the power of the artefact peak observed on *Streaming* recordings decreased as the stimulation frequency increased from 125Hz upwards (1.0mA, 125-140Hz, 60μs).

At high amplitudes in two patients (NL1 and CH5), we also recorded stimulation-related subharmonics. In patient NL1, stimulation of contacts 1L and 2L induced narrow power peaks in the gamma band at half (i.e., 64.9Hz), one-quarter (32.5Hz) and three-quarters (97.4Hz) of the stimulation frequency in the ipsilateral recording only, starting above 2.5mA and stopping abruptly when stimulation amplitude is turned to 0mA, simultaneously with dropping of the 120Hz artefact. (Fig.9) Stimulation of contacts 1R and 2R induced a single power peak at half the stimulation frequency above 3mA and at 2.9mA, respectively (data not shown). The patient had no dyskinesias at the time of the recordings but developed stimulation-related dyskinesia with chronic DBS from contact 2L. At the last follow-up visit, the patient did not show any dyskinesia, but the stimulation-related artefacts were still present (data not shown).

In patient CH5, in a similar setup, only one subharmonic oscillation peak at half the stimulation frequency was recorded (besides the 120Hz artefact), starting from 4mA, and only when stimulating through contact 1R (sensing pair 0-2R). The patient did not display dyskinesia during the recordings (Supplementary Fig.2).

To investigate the nature of these artefacts, we replicated the experiment in a saline setup and recorded no subharmonic oscillations (Fig.9). We cannot exclude a biological nature for these stimulation-related signals.

**Figure 9.**
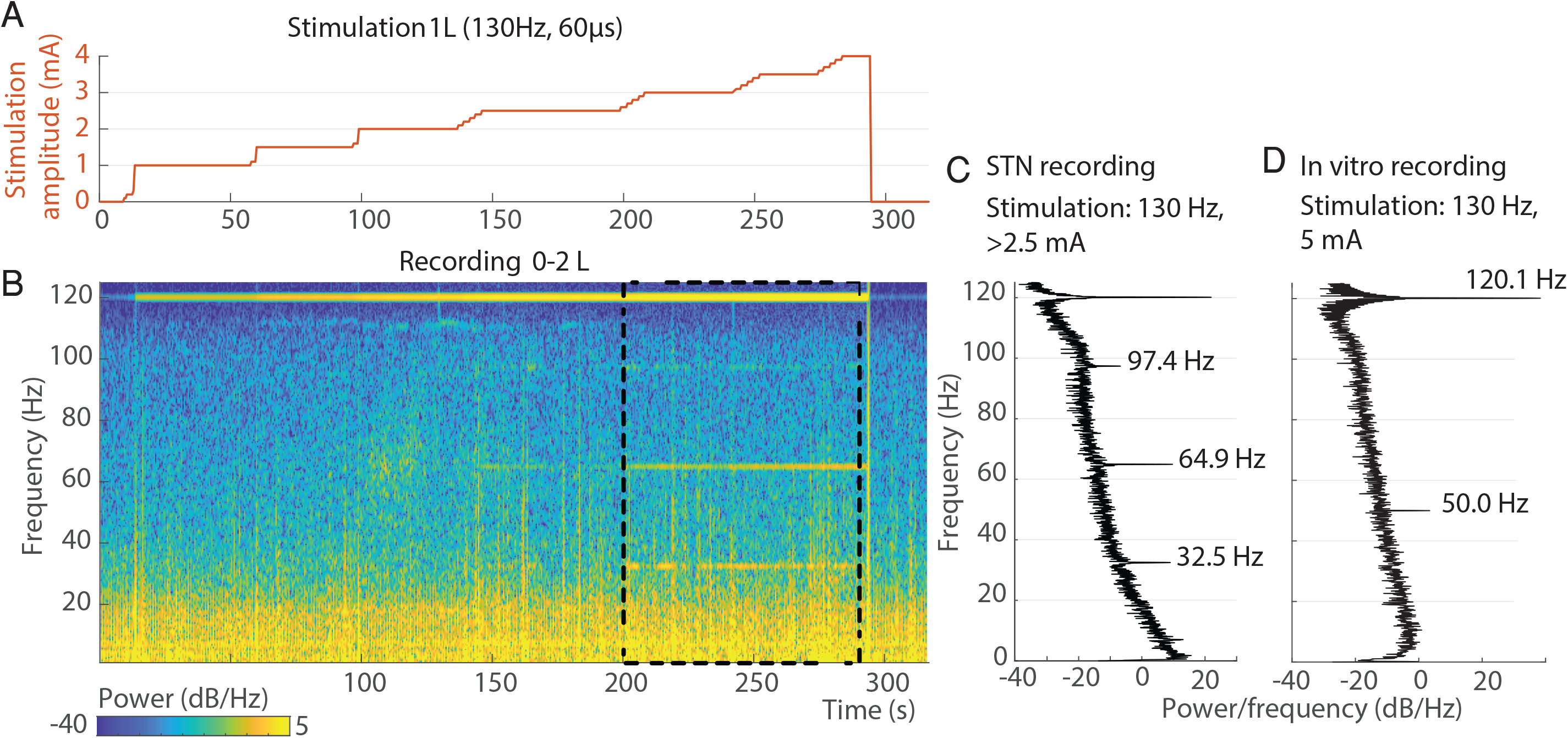
Subharmonic artefacts appearance above a stimulation amplitude threshold. Aligned stimulation amplitude profile **(A)**, local field potential (LFP) spectrogram (B), and power spectrum density (PSD) estimate **(C)** (computed between 200s and 290s) recorded by contact pair 0-2L during monopolar contact review of contact 1L at 130Hz, 60μs and with a stepwise increment of stimulation amplitude from 0mA to 4mA, off-medication (patient NL1). Stimulation at amplitudes of 2.5mA and above induced peak power activities at 35.5Hz, 64.9Hz, and 97.4Hz, which abruptly stopped when stimulation amplitude is lowered to 0mA. The 125Hz-symmetrical artefact at 120Hz is also visible already from 1mA, and is already present with the stimulator turned on at 0mA. No dyskinesias were observed during the recording. Similar findings were recorded when stimulating with contacts 2L, 1R, and 2R, and recording from 1-3L, 0-2R, and 1-3R respectively (data not shown). **(D)** We stimulated saline water at 5mA, 130Hz and 60μs, while recording in *Streaming* mode. Only 50Hz (electrical line power) and 120Hz (stimulation artefact) power bands are visible in this setup.

##### Cardiac-related artefacts

Cardiac artefacts notably affected the power of the beta range. All raw LFP recordings were visually inspected for cardiac-related artefacts. Overall, such artefacts were observed in at least one contact pair in four patients (20%) (Supplementary Table 2).

We observed two categories of cardiac artefact:

1. In three patients (NL1, NL2, and PW1), we observed cardiac artefacts in *Streaming* or *Setup* modes when stimulation was on (even at 0mA) but not when stimulation was off. The artefacts were absent in *Survey* mode. In our experiments, monopolar impedances of the artefactual and stimulation contacts were all in acceptable range (between 785-1643Ω) and we did not observe an imbalance of impedance between an artefactual recording contact and its corresponding stimulation contact (difference range 9-495Ω) (Supplementary. Table 2). The manufacturer suggested that these artefacts may be linked to the involvement of the stimulation contact in the sensing circuitry that might arise from fluid leakage, typically at the leads-extensor connector. The cleaning process effectively removed the QRS peaks from the raw signal, preserving the signal neural content. (Fig.10)
2. In one patient (CH6), we observed cardiac-related artefacts with *Survey Indefinite Streaming, Setup* and/or *Streaming* mode recordings (stimulation “off” and “on”).

**Figure 10.**
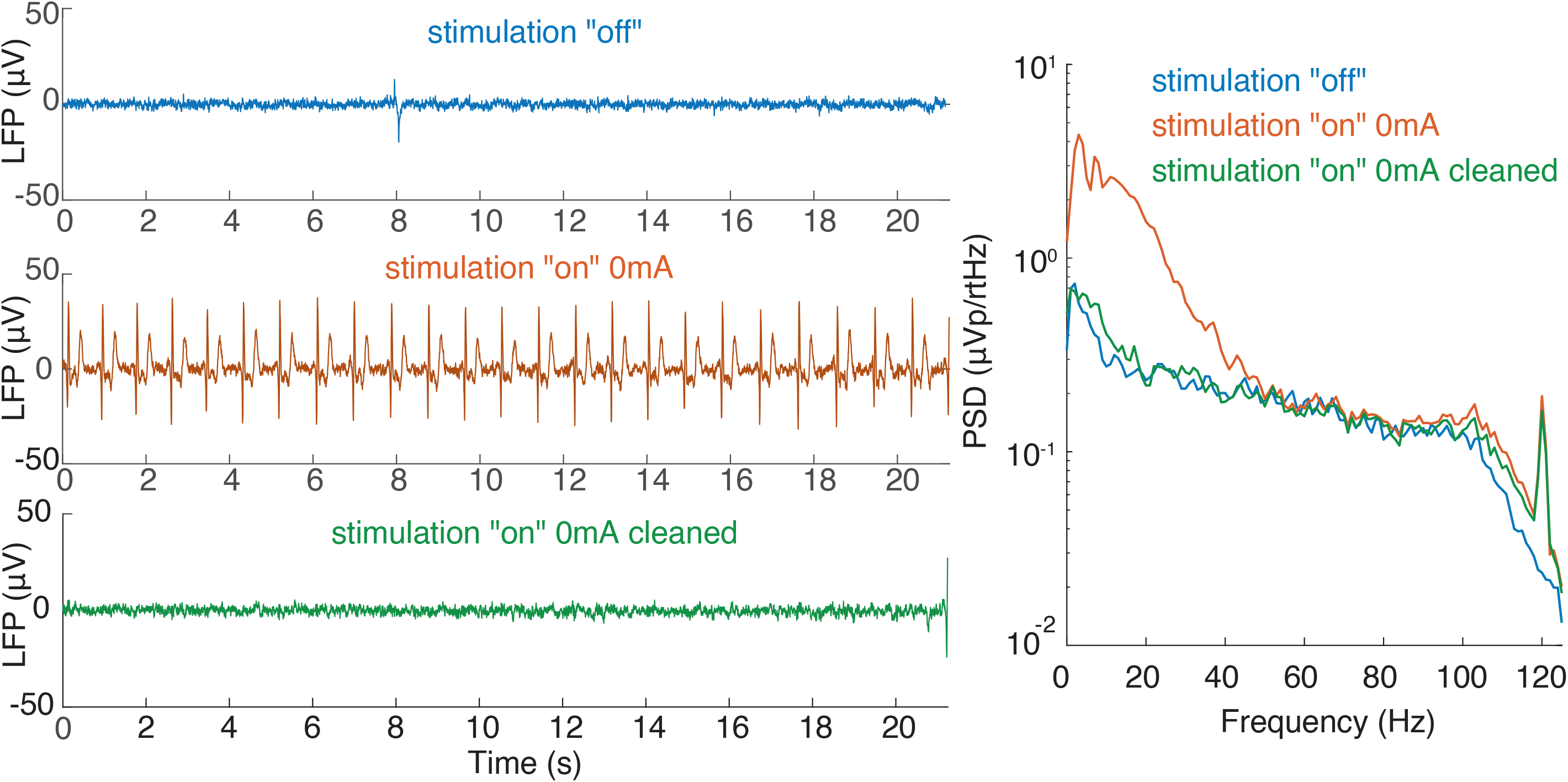
Cardiac artefacts. Illustrative example of local field potential (LFP) signals recorded with *Setup* in the stimulation “off” and stimulation “on” (0mA) conditions (patient NL1, contact pair 1-3R) in the early postoperative period. The inclusion of the active contact (monopolar configuration, referenced to the implantable pulse generator case), even at 0mA, immediately induced cardiac-related artefacts that corrupted the raw signal and covered most frequencies under 50Hz in the power spectrum density (PSD) estimate. The cleaning process effectively removed the QRS peaks from the raw signal. The resulting PSD shows a decrease in the frequency content between 1-40Hz, the range typically affected by the cardiac artefact. The similarity between the PSD in stim “off” and “on” conditions suggests that the neural content of the signal was mainly preserved.

##### Movement-related artefacts

There was an evident gait-related movement artefact in the right hemisphere recordings of PW4 while recording in *Streaming* mode “off” stimulation. (Fig.5). Its presence in one hemisphere only and the lack of relation with the corresponding stepping leg supports the hypothesis of the artefactual origin of these oscillations, rather than a gait-related neural modulation. The IPG was placed in the right abdomen. No cardiac-related artefact was present in these recordings. Impedances were within the normal range for all contact pairs. This artefact’s origin remains unclear. Of relevance, such an artefact was visible exclusively when recordings were epoched to specific events of the gait cycle, i.e., heel strike. We cannot exclude the presence of artefacts during other motor tasks, and LFP recordings with Percept PC should thus be carefully evaluated.

Spike-like artefacts were also noticed in recordings identified as non-artefactual by the Percept PC (Supplementary Fig.1B). This is possibly related to the transient and episodic nature of dystonic and myoclonic jerks, which might be variably captured by the Percept PC. Longer recordings (with *Survey Indefinite Streaming* and *Streaming*) might be more robust against movement artefacts for PSD computation, especially when estimating theta activity, and might facilitate proper contact selection for chronic sensing (Supplementary Fig.3).

### Synchronization of the Percept PC with other devices

Synchronization input/output signals are not presently available within Percept PC. We tested the use of electrical artefact induced by the DBS in the *Survey Indefinite Streaming* and in the *Streaming* mode; alignment may only be performed offline.

DBS through the Percept PC itself can be used to generate in the LFP a stimulation-induced artefact, which may simultaneously be picked up by other external devices (Fig.11). We successfully synchronized the LFP signal measured with Percept PC with signals recorded by EEG and EMG by aligning the stimulation artefact.

We confirmed in saline setup that these artefacts occurred at the time of the first and last pulse (Fig.11C). Their amplitude correlated with the stimulation amplitude (Fig.11D).

**Figure 11.**
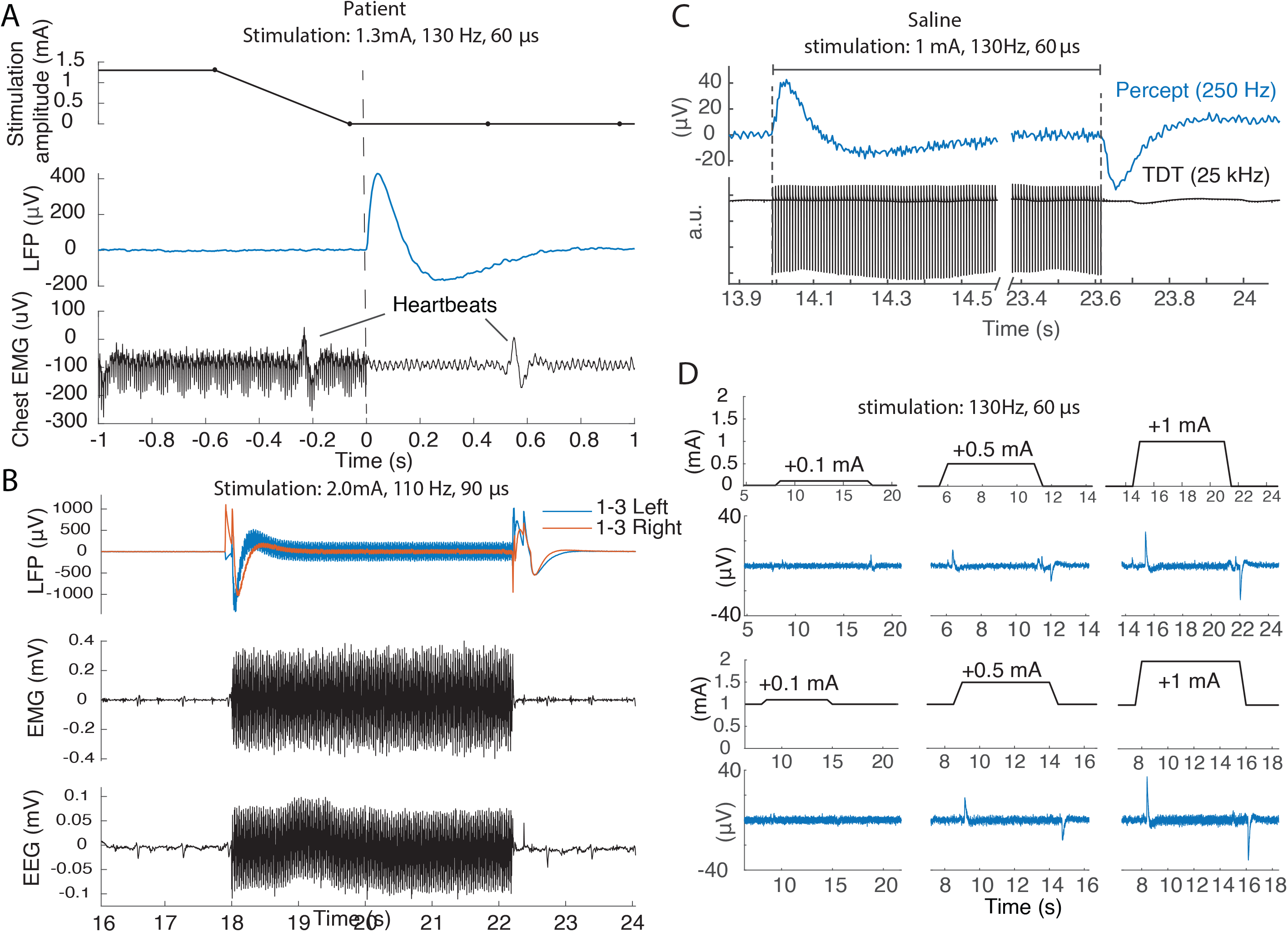
Deep brain stimulation (DBS) artefacts can be used to synchronize recordings in the *Streaming* modality with electromyography (EMG) and electroencephalogram (EEG) recordings. **(A)** A change in DBS amplitude (from 1.3mA to 0mA, 130Hz, 60µs - top) induced an artefact recorded by BrainSense™ *Streaming* (middle) in patient CH5. In addition, a bipolar surface EMG probe located on the chest around the implantable pulse generator (IPG) (one electrode on the clavicle, the other on the IPG - bottom) captured single-pulse artefacts. Therefore, *Streaming* and EMG recordings could easily be aligned (vertical dashed line). Note the temporal inaccuracy of the stimulation amplitude stored at 2Hz (top). (B) Recordings from the two local field potential (LFP) channels (left and right), one EMG channel, and EEG channel FT10 while bilateral stimulation (2.0mA, 110Hz, 90µs) was turned “on” and “off” in BrainSense *Streaming* mode in patient PW6. A ∼0.5s-long transition artefact appeared on the LFP recording when stimulation was turned on (or when amplitude was increased) and, with opposite polarity, when the stimulation was switched off (or when amplitude was reduced). For stimulation frequencies below the Nyquist frequency (125Hz), the pulses could be directly detected on the LFP recordings and added their shape to the onset artefact. Both EMG and EEG clearly show stimulation-related bursts as DBS was turned on. Synchronization could then be achieved offline by aligning the signal to the sharp drop of the stimulation artefact. (C) In saline water, the stimulation amplitude was changed from 0mA to 1mA and from 1mA to 0mA (130Hz, 60µs) while simultaneously recording with *Streaming* and a synchronized high-resolution amplifier. The first and last DBS pulses captured by the amplifier were aligned to the onset of the deflections observed in *Streaming*. (D) In saline comparisons in the amplitude of DBS-induced artefacts for stimulation changes of different amplitudes (+ 0.1mA, +0.5mA, or +1.0mA), either when starting at 0mA (top row) or 1mA (bottom row), confirm that artefacts may be captured independently of the base level of stimulation.

## DISCUSSION

The sensing capabilities of Percept PC open new opportunities to optimize the clinical efficacy of DBS. First, the possibility of monitoring LFP power in chronically-implanted patients, both in-clinic and at home, allows precise readouts of symptom-specific brain activity patterns, their response to therapies, and fluctuations over time. These neurophysiological maps will complement anatomical model-based approaches for DBS programming, used to predict the shape of the volume of tissue activated^23,24^, to support informed stimulation programming. Second, real-time sensing algorithms provide the substrate for novel adaptive DBS systems operating by modulating stimulation parameters in response to an input signal that can represent symptoms, motor activity, or other behavioral features. These new protocols promise truly personalized treatment adherent to everyday life necessities, reducing side-effects and battery consumption, and overall improving therapeutic efficacy for a wider set of motor and non-motor symptoms.

Our results confirm that Percept PC is capable of recording and monitoring the most clinically-relevant biomarkers for PD and dystonia. In PD patients, a beta peak could be identified in at least one contact in 86% of STN (Fig.1, Fig.3) and *Streaming* mode recordings could help clinicians to set the optimal stimulation amplitude for best beta-band power reduction (Fig.1, Supp Fig.2). Percept PC also allows recording gamma-band modulations, opening the quest for new symptom-specific and network-related biomarkers^25-27^, but also defining new challenges (e.g., stimulation-related harmonics, artefacts – Fig.9).

Of relevance, similar stimulation-related signals were recorded earlier in patients implanted with the Activa PC+S^27^. The authors interpreted them as an entrainment of the local gamma activity at half of the stimulation frequency in the presence of dyskinesia. In another study, finely-tuned gamma oscillations were recorded in LFP at half the stimulation frequency in the absence of dyskinesia. Contrary to what we observed, these signals outlasted the stimulation artefact, arguing against an artefactual nature^28^. Although Percept PC is mainly intended for beta- and gamma-band recordings, a theta band was identified in 83% of non-artefactual contacts of dystonic patients, potentially offering a more meaningful approach for DBS programming in these patients.

As a caveat, the relatively low sampling frequency (250Hz) imposes some constraints to the computation of high-frequency oscillations^29^.

The simplicity of manipulation and the signal quality of Percept PC makes it an easy-to-use and reliable tool that can help guide the selection of optimal therapy parameters (e.g., stimulation contact pairs and amplitudes) during in-clinic visits, and monitor the condition of patients over longer periods at home and during daily activities. However, important aspects need to be considered to ensure clean recordings. The most critical aspects were related to contact selection, artefact detection, and data loss.

The choice of the appropriate sensing mode is pivotal to how these aspects are handled by the device. Each mode provides specific pros and cons that make it more or less appropriate depending on the aim of the recording. For example, the *Survey* and *Setup* modes allow short recordings from all and stimulation-compatible contact pairs, respectively. They both check for the presence of artefacts, but only the latter performs this evaluation in simulation “off” and “on” conditions. With the *Streaming* mode, sensing and stimulation contacts are restricted to predefined combinations that greatly reduce the available choices, but with the advantage of recordings of indefinite length simultaneous to active stimulation. To enable sensing, stimulation must be restricted to the middle contact points, which may not be the most clinically effective. Novel segmented electrodes might ease this issue. The *Indefinite streaming* mode allows recordings of indefinite length from all stimulation-compatible contact pairs, but data are not displayed in real time. Also, recordings in stimulation “on” mode are not possible.

The power band of cardiac artefacts overlaps with the beta frequency range and makes them particularly troublesome for proper monitoring of biomarkers, whether in-clinic or at home. Surgical aspects (e.g., using two sutures to seal the connector between lead and extension or selecting the right implant site of the IPG^10,30^) may help to minimize artefacts and need to be carefully considered and confirmed.

The *Timeline* mode provides the long-awaited possibility of chronically recording LFP; however recordings are restricted to the average power of a narrow band (5Hz) around a predefined frequency. This selection remains static for the full extent of the recordings, and may therefore fail to capture modulations or frequency shifts^18^. The possibility of patients manually marking events raises opportunities to obtain precise monitoring of specific symptom manifestations in a real-life environment. While this may be indeed the case for episodes with slow-changing dynamics (minutes to hours), such as off periods, medication-induced dyskinesias, or sleep (Fig.6), its utility for short events such as freezing of gait or falls may be more problematic. Indeed, apart from heavily relying on patients compliance, the inevitable delay between the occurrence of the event and the manual marking with the patient controller, and the fact that only LFP after the marker are recorded would make it difficult to correlate the brain signal related to (the onset of) a short event. Furthermore, in *Timeline* mode the provided power averaged over 10min would most likely miss short neural signatures related to these events^14^.

Complementary to the clinical use, the sensing capacities offered by the Percept PC promises to foster translational and clinical research which, until recently, was limited to a few centers using externalized electrodes or who have access to research-dedicated devices^11,31^. This represents an important springboard for extensive collaborations that aim to identify novel and more specific biomarkers and their true biological meaning.

The *Survey Indefinite Streaming* is the only modality that allows continuous and indefinite LFP measurements with all contact pairs. However, the data streaming cannot be monitored online on the tablet or exported to third-party devices. This critically restricts any real-time application, and renders the correction or optimization of experimental setups inflexible. In this context, synchronization with other devices (e.g., EEG, EMG, etc.) is an important issue also due to the inability to check a successful synchronization artefact online and the lack of an embedded synchronization method; this may represent a relevant limitation for research.

It is important to note that although some LFP data and artefact information is available online and readily accessible through the User Interface, more advanced analysis can only be performed offline.

Data export is industrious and requires dedicated software; the exact time of saving the recording needs to be noted and files need to be monitored for data loss. To simplify this process, we provide an open-source code jointly with this report, which automatically extracts JSON files of the Percept PC and helps to account for missing data.

We hope that many of the above-mentioned drawbacks will soon be addressed and optimized. In the meantime, based on our initial multicenter experience in different clinical and research settings in patients with different diagnoses and in different conditions, we share our practical tips to maximize the performance and signal quality of this novel device (Table 3).

Sensing of LFP-based biomarkers will become routine in clinical practice, paving the way for better understanding and monitoring of distinctive neural signatures of specific symptoms or behaviors. This is a necessary premise for true patient-tailored neuromodulatory therapeutic interventions.

## Data Availability

Data collected in the three centers are available from the respective senior authors at a reasonable request. The software is available at github.com

https://github.com/YohannThenaisie/PerceptToolbox.git

## Acknowledgements

The authors would like to thank Prof. C. Matthies (UKW) for the neurosurgical information, Dr Henri Lorach (CHUV) for their help with data acquisition, and Dr. Scott Stanslaski and Dr. Gaetano Leogrande (Medtronic) for sharing some technical information on the system. The draft manuscript was edited for English language by Deborah Nock (Medical WriteAway, Norwich, UK).

## Credit authorship contribution statement

YT: Conceptualization; Data curation; Formal analysis; Methodology; Software; Visualization; Writing - original draft.

CP: Conceptualization; Data curation; Formal analysis; Methodology; Software; Visualization; Writing - original draft.

AC: Formal analysis; Methodology; Writing - review & editing.

BJK: Formal analysis; Software; Visualization; Writing - review & editing.

PC: Data curation; Supervision; Writing - review & editing.

MCJ: Data curation; Writing - review & editing.

JFB: Data curation; Writing - review & editing.

EM: Data acquisition; Methodology; Writing - review & editing.

LB: Data acquisition; Methodology; Writing - review & editing.

RZ: Data curation; Supervision; Writing - review & editing.

GC: Supervision; Resources; Writing - review & editing.

JB: Data curation; Supervision; Writing - review & editing.

NAvdG: Data curation; Supervision; Writing - review & editing.

CFH: Data curation; Supervision; Writing - review & editing.

EMM: Conceptualization; Data acquisition; Data curation; Formal analysis; Methodology; Resources; Supervision; Writing - original draft; Writing - review & editing.

IUI: Conceptualization; Data acquisition; Data curation; Formal analysis; Methodology; Resources; Supervision; Writing - original draft; Writing - review & editing.

MFC: Conceptualization; Data acquisition; Data curation; Formal analysis; Methodology; Resources; Supervision; Writing - original draft; Writing - review & editing.

