## Supplementary figures and images for "Towards adaptive deep brain stimulation: clinical and technical notes on a novel commercial device for chronic brain sensing"

### Supplementary Figure 1

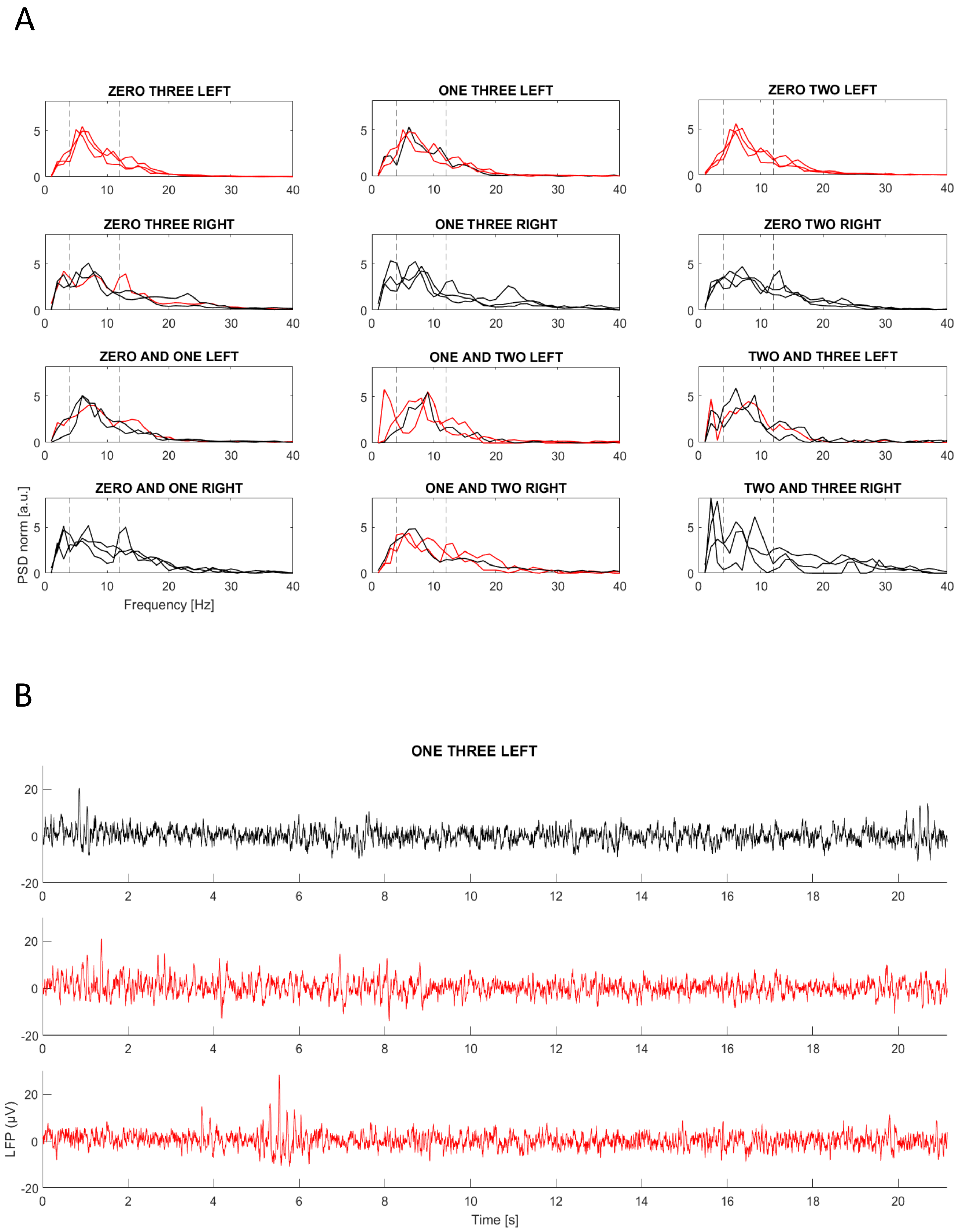

### Supplementary Figure 2

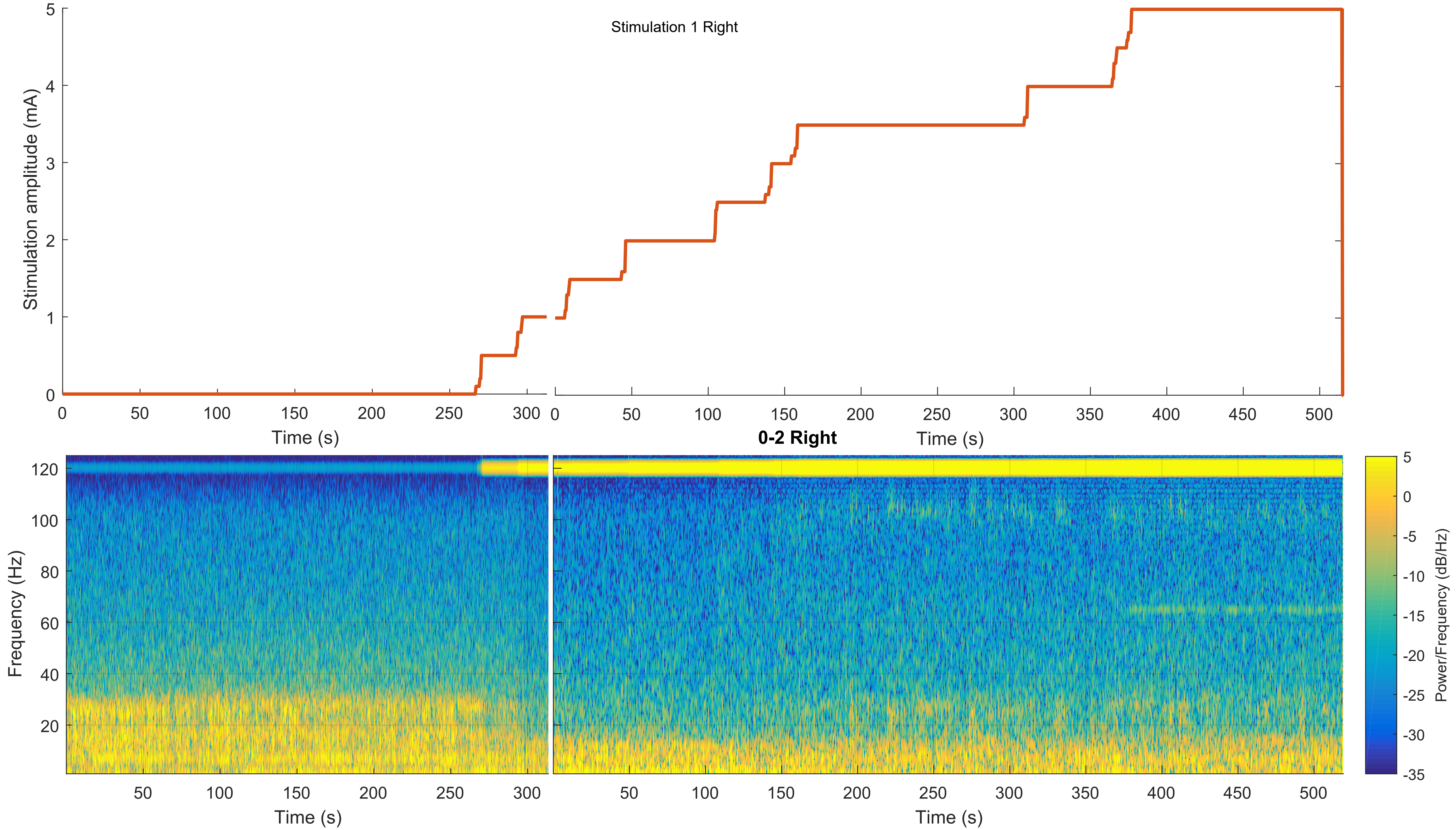

### Supplementary Figure 3

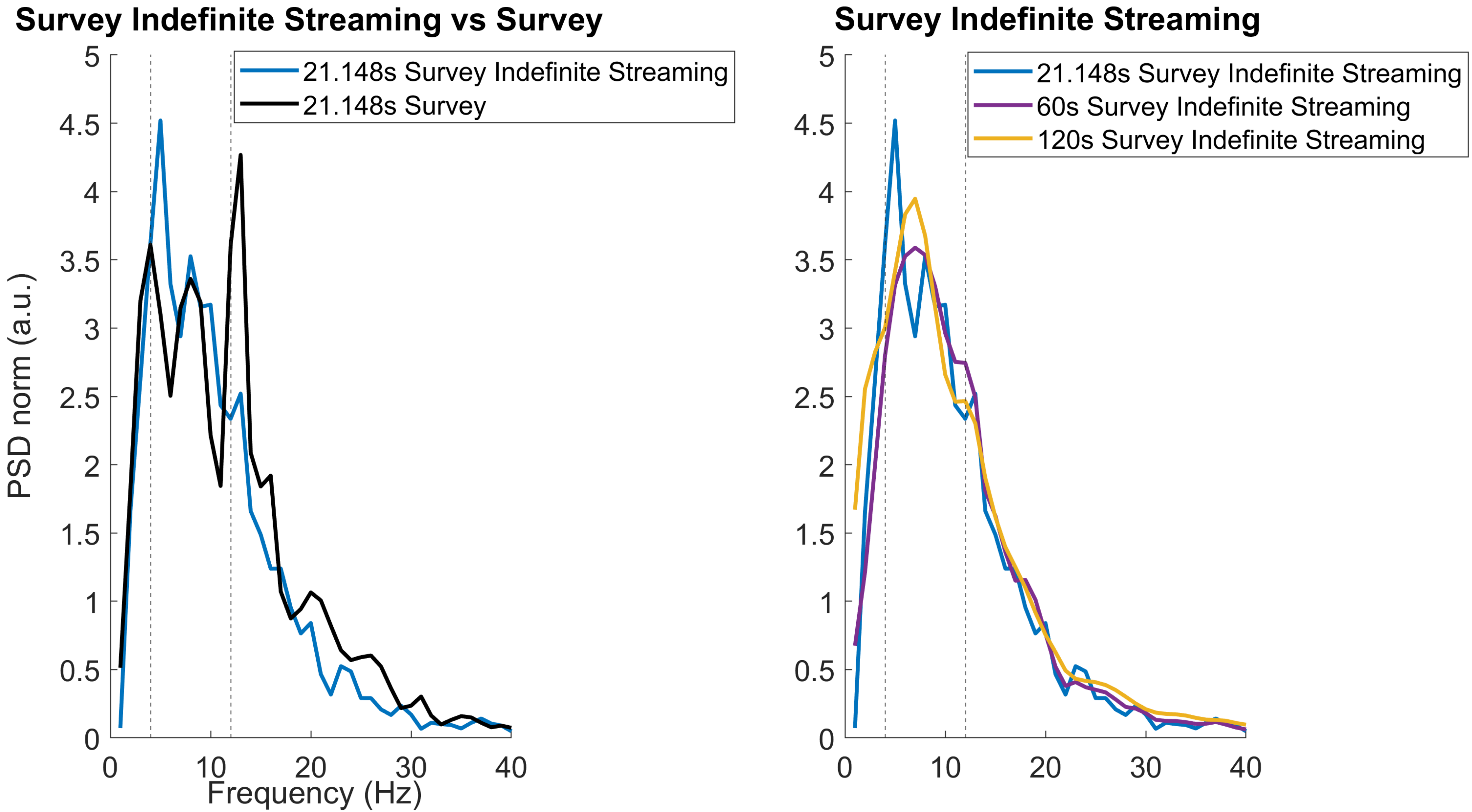
