## Supplementary File 1 text for "Towards adaptive deep brain stimulation: clinical and technical notes on a novel commercial device for chronic brain sensing"

### Supplementary text

#### Surgical procedure

Four patients received the Percept™ PC during replacement of an implantable pulse generator (IPG) and two patients (CH2, CH4) received it five days after lead implantation. All other patients received the Percept PC simultaneously to lead implant. In one patient (PW4), the IPG was implanted in the right abdominal region; in all other patients, it was implanted in the chest (left: NL1, NL2, PW2-3; right: CH1-10, PW1, 5-8, and PW1A).

All patients but CH10 received the deep brain stimulation (DBS) lead model 3389 (Medtronic PLC, USA) with four cylindrical contacts of 1.5mm each and contact-to-contact separation of 0.5mm (contacts 0/8 were the lowermost, whereas contacts 3/11 were the uppermost; for purpose of clarity, we subsequently named them 0-3L and 0-3R). In patient CH10 (with refractory chronic pain in the left hand), the Percept PC was connected with two electrocorticography (ECoG) strips (Resume II, four contacts per lead) for epidural right motor cortex stimulation.

The surgical procedure of each center has previously been reported[1-4]. The proper lead placement in the subthalamic nucleus (STN) for Parkinson's disease (PD) patients and within the globus pallidus pars interna (GPi) region for dystonic patients was checked by means of intraoperative microelectrode recordings and test stimulation, and by image fusion of pre- and postoperative scans. No specific procedures were followed for the Percept PC implants. Of note, we used a standard silicon sleeve (Medtronic PLC, USA) to cover the connection between the lead and extension cables, and fixed it with one stich on the distal end (Haga/LUMC) or two stitches at the proximal and distal end (CHUV, UKW). The lead was fixed to the skull with a silicon cap (StimLoc, Medtronic, PLC) (UKW, LUMC) or with cement (Palacos R+G, Herserus Medical, Germany) (CHUV).

#### Patient recordings

The timing of recordings, medication and stimulation state, and recording modality used are reported in Table 1. For all CHUV subjects, we collected local field potentials (LFP) for at least 1 min in the eyes-open resting state, using the *Survey Indefinite Streaming* mode (CH1-4, 6-9) or the *Streaming* mode (CH5). Additionally, we recorded modulations during a motor task (i.e., knee extension movements while sitting) in the *Survey Indefinite Streaming* mode (CH 1-4) or the *Streaming* mode (CH5-9 – Fig.2C).

For all UKW subjects, we collected LFP during the eyes-open resting state. LFP of patient PW4 were also acquired during unperturbed walking with *Streaming* mode in a gait laboratory environment[3,5,6]. Recordings in PD patients were performed after pausing the stimulation for at least 30 minutes. In one case (PW1, Fig.2B), we also performed the recordings in meds "on" and 30 minutes after switching on the stimulation at the clinically effective parameters. In patients with dystonia, we aimed for a longer pause of DBS (72 h for PW4-5-1A-8 and 12 h for PW6). These patients were not taking medication during the recordings.

For Haga/LUMC subjects, recordings were obtained in the context of routine monopolar contact review, outpatient evaluations, and at home (*Timeline).*

***In vitro* recordings**

Technical tests performed in saline water were run at CHUV. A DBS lead (model 3389, Medtronic, USA) was inserted in a saline bath and simultaneously connected to the Percept PC and to a high-resolution external amplifier able to record at 24414.06Hz (RZ5D, Tucker Davis Technologies, TDT, USA). Signals from both systems were synchronized by sending an external 10Hz sinusoidal signal generated with Agilent 33210A LXI at 100mA for a few seconds. Tests aimed to:(i) validate the nominal sampling frequency *Fs* of the Percept PC in the presence of a pure sinusoidal signal generated with a high-precision function generator, computed as *Fs=(number of samples)/(number of oscillations)*(frequency of oscillations)*; (ii) evaluate the impact of stimulation artifacts; (iii) validate synchronization methods.

**Analysis of recordings in PD patients**

Beta-band analysis was performed on *Survey Indefinite Streaming* (CHUV) or *Survey* (UKW, Haga-LUMC; Supplementary Table 1, Fig.3). For each *Survey Indefinite Streaming*, we reconstructed bipolar LFP signals from adjacent contacts by subtracting the signals extracted from the JavaScript Object Notation (JSON) file as: $LFP_{0\text{-}1} = LFP_{0\text{-}3} - LFP_{1\text{-}3}$, $LFP_{1\text{-}2} = LFP_{1\text{-}3} + LFP_{0\text{-}2} - LFP_{0\text{-}3}$, $LFP_{2\text{-}3} = LFP_{0\text{-}3} - LFP_{0\text{-}2}$. Power spectrum density (PSD) estimates were computed for each hemisphere via Welch's method (*pwelch* function). We defined the frequency of the beta peak $f_{Peak}$ as the frequency with the maximum local maxima power in the 13-35Hz range. We visually verified each PSD and excluded contact pairs without local maxima in the beta band. For each STN, we reported the contact pair with the highest $f_{Peak}$ power.

Short-time Fourier transform was applied on raw LFP from all *Streaming* recordings (Table 1). We visually inspected the presence of gamma band (60-90Hz) in all resulting PSD and spectrograms.

**Analysis of the recordings in dystonic patients**

PSD were computed with Welch's method and 1/f component removal[7]. Each PSD was normalized for the standard deviation computed between 6-96Hz to allow comparison across trials and patients[8]. Artefact-free recordings as identified by the Percept PC were visually inspected for peaks in the theta (4-12Hz) band[8] (Fig.4) and for movement artefacts (i.e., dystonic muscle contractions, tremor and myoclonic jerks) across repetitions. (Supplementary Fig.1) LFP recordings during gait (PW4) were recorded and synchronized with the kinematic data. (Fig.5) The patient was asked to stand quietly and to start walking after a verbal cue over an 8m long walkway. The task was repeated four times. Body kinematics were measured with a full-body marker set and a motion capture system (SMART-DX, BTS, Italy)[9,10]. LFP data were epoched in 800 ms windows centered at the heel contacts. In total, 37 epochs of gait were analyzed.

##### Comparison of recordings

The company informed us that to compute PSD, Percept PC first scales data to micro-Volts peak by multiplying raw LFP by 1.60. PSD is then computed via Welch's method (1s-windows, 0.6s overlap, 1Hz frequency resolution).

This information is relevant for example when comparing an FFT calculated by Percept PC (in uVp) with an FFT calculated offline starting from raw data using Welch's method. For all signals shown in the user interface, the correction (when needed) is already performed.
