## Supplementary File 2 Sup fig captions for "Towards adaptive deep brain stimulation: clinical and technical notes on a novel commercial device for chronic brain sensing"

### Supplementary figure captions

**Supplementary Figure 1. Results from three subsequent *Survey* recordings in one dystonic patient (PW5)**

**(A)** Power spectrum densities (PSD) of each recording and contact pair. Vertical dashed grey lines represent the theta band (4-12Hz). In red, the recordings labeled as artefactual by the Precept™ PC; in black, the non-artefactual. **(B)** Raw data of three *Survey* recordings from the electrodes pair 1-3L. Of note, transient movement artefacts are present in all recordings (also in the one labeled non-artefactual).

**Supplementary Figure 2. Titration of stimulation**

In patient CH5,stimulation (130Hz, 60µs) was applied at contact 1R while recording was performed with *Streaming* between contacts 0-2R. A high power was elicited at 65Hz while stimulating above 4mA. The stimulation artefact at 120Hz is evident from 0.5mA, but is already present with the stimulator turned on at 0mA. Suppression of the beta band recorded off stimulation is apparent starting at 1mA. Note that the recording was paused and resumed (white vertical bar).

**Supplementary Figure 3. Power spectrum density (PSD) analysis of one dystonic patient (PW5) in the resting state**

Data were recorded from a non-artefactual contact pair (0-2R) for different durations and with different recording modalities during resting. Vertical grey dashed lines represent the theta band.

**(A**) Comparison between PSD estimates of *Survey Indefinite Streaming* and *Survey* local field potential (LFP) data over a window of 21.148s (default *Survey* duration). The two spectral representations provided similar peaks in the theta band but both showed a jagged profile. Such a short window duration might be not robust to movement artefacts for a reliable PSD estimation.

**(B)** Comparison between PSD estimates of *Survey Indefinite Streaming* LFP data computed over different window durations (i.e., 21.148 s, 60 s, 120 s). The averaging over more windows provided a smoother PSD, possibly more trustworthy and robust to the influence of episodic artefacts.
