## Supplementary Table 1 for "Towards adaptive deep brain stimulation: clinical and technical notes on a novel commercial device for chronic brain sensing"

**Supplementary Table 1. Location of maximum beta peak power and best clinical contact on Parkinson's disease patients**

| | | Indefinite streaming at rest | | Stimulation with best clinical outcome | |
|---|---|---|---|---|---|
| **Subject** | **Side** | **Contact pairs with highest beta power** | **Recording[a]** | **Stimulation contact** | **Stimulation parameters** |
| CH1 | L | 0-2 | IS (69s) | 1- | 1.7mA, 130Hz, 60µs |
| | R | 0-2 | | 1- | 1.6mA, 130Hz, 60µs |
| CH2 | L | 1-3 | IS (128s) | 0- | 1.7mA, 130Hz, 60µs |
| | R | 1-3 | | 1- | 1.6mA, 130Hz, 60µs |
| CH3 | L | 0-3 | IS (121s) | 3- | 3.0mA, 130Hz, 60µs |
| | R | 0-3 | | 3- | 1.9mA, 130Hz, 60µs |
| CH4 | L | 1-3 | IS (113s) | 2-3- IL[b] | 2.0mA, 180Hz, 90µs |
| | R | No beta[d] | | 2-3- IL[b] | 2.1mA, 180Hz, 90µs |
| CH6 | L | 1-2 | IS (90s) | 1- | 1.4mA,130Hz, 60µs |
| | R | 1-3 | | 1- | 1.4mA, 130Hz, 60µs |
| CH7 | L | 0-3 | IS (107s) | 1- | 4.7mA, 140Hz, 90µs |
| | R | 0-3 | | 3- | 2.9mA, 140Hz, 90µs |
| CH8 | L | 0-3 | IS (105s) | 2- | 1.0mA, 245Hz, 60µs |
| | R | 0-2 | | 3- | 3.2mA, 245Hz, 60µs |
| CH9 | L | 1-2 | IS (111s) | 3- | 2.0mA, 130Hz, 60µs |
| | R | 1-3 | | 2- | 2.0mA, 130Hz, 60µs |
| NL1[C] | L | 1-3 | Survey | 1- | 4.3mA, 130Hz, 60µs |
| | R | 2-3 | | 3- | 1.2mA, 130Hz, 60µs |
| NL2 | L | No beta[d] | Survey | 0-1+ | 2.8mA, 185Hz, 60µs |
| | R | No beta[d] | | 1+2-3- | 3.0mA, 185Hz, 60µs, |
| PW1[C] | L | 0-3 | Survey | 2+3- | 5.0 mA, 180Hz, 60µs |
| | R | 1-3 | | 2- | 2.6mA, 180Hz, 60µs |

[a] Data from 21 s of *Survey* or from a variable duration of *Survey Indefinite Streaming* (IS) recordings.
[b] Interleaved stimulation (IL).
[c] *Survey* were recorded on four (NL1) or two (PW1) sessions. Only the recording exhibiting the clearest beta peaks was considered for analysis.
[d] In three STN (two patients), no clear beta peak could be identified in any of the contact pairs.
