## Supplementary Table 2 for "Towards adaptive deep brain stimulation: clinical and technical notes on a novel commercial device for chronic brain sensing"

### Supplementary Table 2: Patients with cardiac artefacts

<table>
<tr><th>Patient</th><th>IPG implant site</th><th colspan="2">Modes</th><th>Artefactual channel(s)</th><th>Monopolar contact impedance</th></tr>
<tr><td rowspan="2">NL1</td><td rowspan="2">Chest left</td><td colspan="2">Streaming and Setup<br>Stimulation “on” (left or right)<br><br>Stimulation “off”</td><td>0-2L<br>1-3R<br><br>—</td><td rowspan="2">0L: 967 Ω<br>1L: 1070 Ω (Stim)<br>2L: 1126 Ω<br>1R: 971 Ω<br>2R: 962 Ω (Stim)<br>3R: 1006 Ω</td></tr>
<tr><td colspan="2">Setup*</td><td>All contact pairs</td></tr>
<tr><td rowspan="3">NL2</td><td rowspan="3">Chest left</td><td>Session 1</td><td>Streaming<br>Stimulation “on” (left or right)</td><td>0-3L<br>1-3L<br>0-2R<br>0-3R</td><td>0R: 1224 Ω<br>1R: 1409 Ω (Stim or OFF/0mA)<br>2R: 1387 Ω (Stim or OFF/0mA)<br>3R: 1592 Ω</td></tr>
<tr><td rowspan="2">Session 3</td><td>Streaming<br>Stimulation “on” (right)<br><br>Stimulation “off”</td><td>0-3R<br><br>—</td><td rowspan="2">0R: 1643 Ω,<br>1R: 1189 Ω (Stim or OFF/0mA)<br>2R: 1186 Ω (Stim or OFF/0mA)<br>3R: 1685 Ω</td></tr>
<tr><td>Setup*</td><td>All contact pairs</td></tr>
<tr><td>PW1</td><td>Chest right</td><td colspan="2">Streaming<br>Stimulation “on” (left and right)</td><td>1-3L<br>1-3R</td><td>1L: 1046 Ω<br>2L: 1128 Ω (Stim)<br>3L: 1445 Ω<br>1R: 829 Ω<br>2R: 996 (Stim)<br>3R: 815 Ω</td></tr>
<tr><td>CH6</td><td>Chest right</td><td colspan="2">Survey Indefinite Streaming</td><td>1-3R</td><td>1R: 802Ω<br>3R: 785Ω</td></tr>
</table>

| | | | |
|---|---|---|---|
| | | *Setup** | Five out of six contact pairs (including 1-3R) |

* In the Setup mode, contact pairs were labelled as “artefactual” by the Percept system; in the other modalities, artefacts were detected offline by visual inspection.
